# Detection of novel *Plasmodium falciparum* haplotypes under treatment pressure in pediatric severe malaria

**DOI:** 10.1101/2024.08.20.24312296

**Authors:** Balotin Fogang, Emilie Guillochon, Claire Kamaliddin, Gino Agbota, Sem Ezinmegnon, Maroufou Jules Alao, Philippe Deloron, Gwladys Bertin, Antoine Claessens

## Abstract

**Background:** In Africa, the clearance time for *P. falciparum* severe malaria varies significantly, likely due to the complexity of *P. falciparum* infections and the sequestration phenomenon exhibited by this parasite. This study aims to evaluate different methods to study intra-host dynamics of polygenomic infections during parasite clearance under antimalarial treatment. Additionally, it seeks to determine the association between parasite clearance rate following artesunate or quinine treatment and the genetic complexity of *P. falciparum* in Beninese children with severe malaria.

**Methods:** Sixty-five *P. falciparum* severe malaria individuals diagnosed by microscopy and treated with artesunate or quinine were sampled every 8 hours for 24 hours. Using whole genome sequencing (WGS) data, we estimated the multiplicity of infection (MOI) with three algorithms (*Fws*, THE REAL McCOIL, and RoH). We then characterized the *P. falciparum* genetic complexity in WGS-identified polyclonal infections using amplicon sequencing (AmpSeq) on DNA extracted from plasma and from the red blood cells pellet.

**Results:** AmpSeq demonstrated greater sensitivity in detecting multiple genomes within isolates compared to WGS methods. The MOI from AmpSeq was significantly higher in RBC pellets compared to plasma (2.4 vs 1.8 distinct microhaplotypes per isolate). However, at parasitaemia over 1000 parasites/uL, the same MOI was detected in both plasma and pellet samples in 85.4% of the isolates. We observed a high variability in parasite clearance rate among participants, but it was not associated with parasite MOI at diagnostic. Interestingly, in 60.9% of participants, previously undetected microhaplotypes appeared in circulation 16 hours after treatment initiation.

**Conclusion:** These findings demonstrate that combining different haplotyping techniques effectively determines parasite genetic complexity. Additionally, plasma can be effectively used for parasite genotyping at sufficient parasitaemia levels. The parasite clearance rate of severe malaria is independent of parasite MOI. However, genotyping a single blood sample upon hospital admission does not capture the full spectrum of parasite genotypes present in the infection.

## Introduction

*Plasmodium falciparum* is the predominant malaria species and a major contributor to child mortality in Africa. Many children infected with *P. falciparum* succumb to the disease before even reaching a hospital or clinic. Among those who are admitted with severe malaria and receive parenteral antimalarial treatment, approximately 5% do not survive, highlighting the urgent need for effective interventions [1]. Since 2011, the World Health Organisation (WHO) has recommended intravenous artesunate as the first-line antimalarial treatment for severe malaria [2], citing its potential to reduce severe malaria mortality rates by 22.5% compared to quinine [3]. Nevertheless, in cases where artesunate is unavailable, alternative severe malaria treatment such as intramuscular artemether or intravenous quinine are recommended [2].

Despite the widespread recommendation of Artemisinin-based Combination Therapies (ACTs) as the frontline treatment for malaria, reports of *P. falciparum* resistance to ACTs have risen in Asia [4, 5] and more recently in Africa [6, 7]. Indeed, artemisinin resistance is characterized by the slow clearance of parasites *in vivo*, which is a result of reduced drug susceptibility in ring-stage parasites driven by mutations in the *P. falciparum Kelch13* gene [8]. Since the introduction of artemisinin-based combination therapy (ACT) in 2005, no Pfkelch13 mutation associated with artemisinin resistance has been identified in Benin [9, 10].

Although the *PfKelch13* polymorphisms are the strongest predictor of *in vivo* parasite clearance time under treatment with artesunate, this time can be affected by other factors such as the age of the host, host-acquired immunity, initial parasitemia levels, and the developmental stages of parasite [11–13]. Although controversial, studies have shown a link between the genetic complexity of *P. falciparum* and delayed parasite clearance in Africa. Indeed, children infected with multiple strains had nearly a 3-fold increase in treatment failure compared to their age mates infected with a single strain [14]. However, low multiplicity of infection (MOI) at baseline were associated with detectable parasitaemia at 72h post-ACT treatment [15]. The association between antimalarial parasite clearance rate and parasite genetic complexity in individuals with severe malaria (SM) remains to be explored.

In SM, the delay in parasite clearance could also be attributed to the significant sequestration of mature *P. falciparum* parasitized erythrocytes within the tissue capillaries. Mathematical modelling using plasma histidine-rich protein 2 (PfHRP2) levels and parasitaemia have indicated that the number of sequestered parasites is substantially higher than what is detected in the peripheral blood [16–18]. Because the circulating parasitaemia is highly dependent on the parasite developmental stage, the amount of *Plasmodium* DNA in plasma is actually more accurate than parasitemia for diagnostic differentiating severe from uncomplicated malaria [19]. Other studies have also demonstrated that plasma can be used to detect and quantify the *Plasmodium* DNA by PCR [20, 21]. However, whether genomic DNA (gDNA) extracted from plasma could be used as material to reliably estimate the MOI is not known. Although the plasma may contain less parasite DNA than within RBCs, it is unaffected by sequestration and could theoretically contain parasite DNA released from prior schizont ruptures of all the parasite genotypes present within an individual. However, there is currently no evidence regarding the sensitivity of *Plasmodium* genotyping when using plasma samples as the DNA source.

Various sequencing approaches have recently been used to assess the complexity of *P. falciparum*, quantified as the number of parasite genotypes within an isolate [22]. These methods include whole-genome sequencing (WGS)-based techniques using diverse metrics such as *Fws* [23], THE REAL McCOIL [24], and Runs of Homozygosity (RoH) [25], which elucidate intra-host parasite diversity by analysing genome-wide variations across multiple loci. *Fws* is a metric characterizing within-host diversity and its relationship to population-level diversity, with theoretical values ranging from 0 to 1. The RoH indicates genetic relatedness between 2 or more genotypes within an isolate by identifying long blocks of haplotypes that have been inherited from the same parent. The REAL McCOIL is a statistical approach that uses data from all infections to simultaneously estimate allelic frequency and the number of distinct genotypes s in an isolate. Amplicon sequencing (AmpSeq), a technique targeting highly polymorphic loci, has emerged as a novel tool capable of detecting minority microhaplotypes with within-sample frequencies as low as 0.1% [26]. Indeed, the strength of sequencing targeted non-repetitive regions that harbour extensive single nucleotide polymorphisms (SNPs) lies in the fact that all SNPs within an amplicon are linked by a single sequence read, enabling direct microhaplotype identification [26]. Therefore, this study aims to evaluate the association between parasite clearance rate and multiplicity of infection in Beninese children undergoing treatment for severe malaria, using a variety of sequencing methods and biological materials.

## Methods

### Study design

Ethical clearance was obtained from “Comité d’Ethique de la Recherche CER_ISBA Benin” (clearance n°90, 06/06/2016 and clearance n°38, 16/05/2014). During the high malaria transmission season in Southern Benin, children under the age of six diagnosed with severe malaria infection at the “Centre Hospitalier et Universitaire de la Mère et de l’Enfant-Lagune (CHU-MEL) de Cotonou” were enrolled in the study in both 2014 and 2016, as described in Kamaliddin 2019 [27]. Prior to their participation, written informed consent from parents or guardians was obtained. Malaria infection was diagnosed in febrile patients by detecting *P. falciparum* on a Giemsa-stained thick blood smear. Parasite density was estimated by counting the number of parasites per at least 200 white blood cells (WBC) and assuming a standard total blood WBC count of 8000 WBC/µL. Diagnosis and classification of severe malaria were carried out in accordance with WHO guidelines for the management of severe malaria [2]. In brief, cerebral malaria (CM) was characterised by impaired consciousness, indicated by a Blantyre score < 3, after excluding other causes of coma. Severe anaemia (SMA) was identified by a haemoglobin level < 5 g/dL or a haematocrit level < 15%. Severe non-cerebral malaria (SNCM) was defined by the features of severe malaria excluding CM involvement. Furthermore, clinical appreciation was at the discretion of the admitting physician.

Approximately 5 mL of blood were collected from all participants using EDTA-coated tubes before the administration of quinine (in 2014) or artesunate (in 2016) therapy (H0). Subsequently, three additional blood samples were taken at +8 hours (H8), +16 hours (H16), and +24 hours (H24) post-treatment initiation to capture parasite dynamics.

### DNA extraction

Plasma was separated from total blood cells by centrifugation at 1500 rpm for 5 min and immediately stored at −20°C. Red blood cell (RBC) pellet was obtained by depleting white blood cells using a gradient-based separation technique Ficoll (GE Healthcare Life Science). *P. falciparum* genomic DNA (gDNA) was extracted from both 200 µL of red blood cell (RBC) pellets and 200 µL of plasma using the DNEasy Blood kit (QIAGEN) according to the manufacturer’s instructions.

### Complexity of infection using whole genome sequencing (WGS) data

WGS was performed by the Malaria Genomic Epidemiology Network (MalariaGEN) and the samples are part of the *Plasmodium falciparum* Community Project accessible at https://www.malariagen.net/project/p-falciparum-community-project/. The raw reads are available on the ENA server under the accession numbers specified in the Supplementary Table 1. Variant Calling Files (VCF) was generated by MalariaGEN as previously described [28]. Briefly, reads were mapped to *P. falciparum* 3D7 v3 reference genome using bwa mem, and the resulting BAM files were subjected to cleaning using Picard tools and GATK. SNPs and indels were called using GATK HaplotypeCaller, with only the core genome considered for analysis.

For downstream analysis bcftools v1.13 [29] and bedtools v2.30 [30] were used for file manipulation. Samples with at least 50% coverage of the genome at 5x were considered. Fraction of within-sample (*Fws*) metrics were computed using the moimix R package, as previously described (https://github.com/bahlolab/moimix). An *Fws* value < 0.95 was indicative of a polyclonal infection. THE REAL McCOIL categorical method was used to estimate the complexity of the infection (MOI) as described by Chang et al. [24] (https://github.com/Greenhouse-Lab/THEREALMcCOIL). Long runs of homozygosity (RoH) and the analysis of heterozygosity in mixed samples were carried out using a custom Python script based on Pearson et al. [25].

### DNA preparation and amplicon sequencing

Three AmpSeq markers, including the genes for conserved plasmodium membrane protein (*cpmp*, PF3D7_0104100), conserved plasmodium protein (*cpp*, PF3D7_1475800) and apical membrane antigen (*ama1-D3*, PF3D7_1133400) were amplified using nested-PCR, as described in [31]. Briefly, primary PCRs were conducted in multiplex for *cpmp/ama1-D3* and monoplex for *cpp*. Nested PCRs were subsequently performed individually for each marker. All amplifications were carried out using the KAPA HiFi Hot Start Ready Mix (Roche) on an Eppendorf Mastercycler Nexus thermocycler. The quality and quantity of nested-PCR products were assessed through gel electrophoresis. Following this, nested-PCR products from each sample were combined in the following proportions: 11 μL of cpmp, 8 μL of cpp, and 4 μL of ama1-D3, for a total of 25 μL. PCR primers and amplification conditions are shown in the Supplementary Table 2.

Libraries preparations and amplicon sequencing were performed by the Genseq platform of the Labex CeMEB (Montpellier). Merged PCR products were purified, sequencing adapters and multiplexing indices were linked by PCR. Indexed products were purified, pooled and quality was verified by electrophoresis using The Fragment Analyzer system (Agilent Biotechnologies). PCR products resulting from RBC pellet or plasma gDNA were separated in two different plates, to prevent the impact of the difference in DNA concentration on the sequencing quality. Finally, amplicon libraries were sequenced on an Illumina MiSeq system in paired-end mode (2 x 300 cycles, 300 bp). The raw AmpSeq reads are available on the Zenedo (https://zenodo.org/records/13224728), with corresponding individual accession numbers provided in Supplementary Table 3.

Markers haplotypes were reconstructed using HaplotypR R package [26], accessible at https://github.com/lerch-a/HaplotypR. Briefly, the quality of raw reads data was assessed using FastQC. Reads corresponding to each marker were demultiplexed based on primers sequences, with subsequent truncation of primers. Reads were trimmed following sequence quality and then fused. SNPs and microhaplotypes were called with default parameters, requiring a minimum coverage of 3 reads per microhaplotype, at least 25 reads coverage per sample, and a within-host microhaplotype frequency ≥ 0.1%. Microhaplotype sequences that differ by less than two SNPs were deemed identical. The relative frequency of each microhaplotype in an isolate was calculated by dividing the number of reads for each microhaplotype by the total number of reads for that isolate. For downstream analysis, marker showing the highest number of microhaplotypes in an individual was retained. The same marker was chosen for the four time points in an individual. The raw data for AmpSeq are presented in Supplementary Table 3.

### Statistical analysis

All statistical analyses were performed using R software version 4.3.2 [32]. Median comparisons of two quantitative variables were conducted using the Mann–Whitney test, while mean comparisons were assessed using the unpaired t-test. The correlation between two quantitative variables was assessed using the Spearman correlation test. For categorical variables, proportions were analysed using the Chi-squared test. We used linear interpolation to estimate the time required for a 50% reduction in parasitaemia, referred as the extrapolated parasite clearance half-life (PC_50%_). The clearance curve for each microhaplotype (parasite genotype detected by AmpSeq) was calculated by multiplying the parasitaemia at each time point by the relative frequency of each microhaplotype. Shannon entropy was used to characterize the complexity of microhaplotypes within an isolate obtained from AmpSeq.

## Results

### Characteristics of the study population

During the years 2014 and 2016, a total of 65 children aged 0 to 5 years old suffering from severe malaria and admitted at CHU-MEL in Benin were recruited. Of these participants, 17 received quinine in 2014, while 48 received artesunate in 2016 (Figure 1). These individuals were classified as 26 cerebral malaria (CM), 16 severe non-cerebral malaria (SNCM) and 23 severe malaria anaemia (SMA) (Supplementary Table 1). The median age and sex ratios of participants were comparable between quinine and artemisinin treated groups. However, the geometric mean parasitaemia in 2014 was significantly higher than in 2016 (p< 0.0001). The demographic and parasitological characteristics of the participants at enrolment are provided in Table 1. Blood samples were also collected on arrival at hospital and every 8 hours for 24 hours (Figure 1).

**Figure 1.**
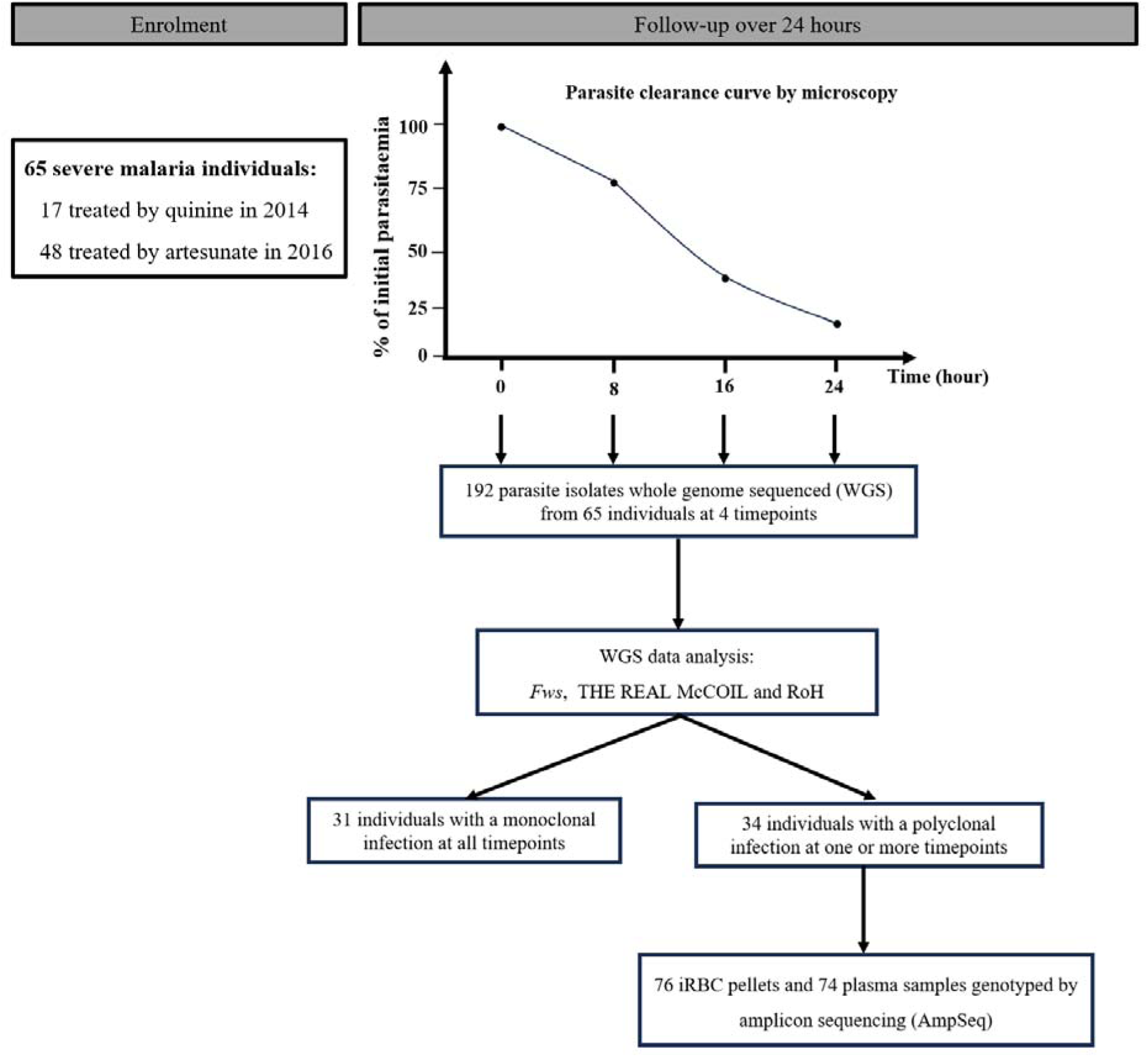
Study flow diagram. Sixty-five severe malaria patients were sampled at the start of treatment and every 8 hours for 24 hours afterwards. Out of 230 isolates submitted for whole genome sequencing, 192 yielded high-quality genomes. The multiplicity of infection (MOI) was determined from parasite whole genome sequences. From 34 individuals with at least one polyclonal timepoint, amplicon sequencing (AmpSeq) was performed on plasma and red blood cell pellet samples. A total of 76 RBC pellet isolates and 74 plasma gDNA isolates from 23 individuals were successfully genotyped.

**Table 1.** Demographic and clinical background of the study participants.

| Characteristics | Artesunate | Quinine | p-value |
| --- | --- | --- | --- |
| Number of participants (N= 65) | 48 | 17 |  |
| Sex ratio (M/F) | 1.9 (27/14)<br>NA= 7 | 1.3 (9/7)<br>NA= 1 | 0.5506 |
| Median age, months (range) | 30 (5-60)<br>NA= 5 | 36 (9-59) | 0.2144 |
| Geometric mean parasitaemia/ μL (range) | 66595<br>(405-2153565) | 1317219<br>(15375-3200000) | < 0.0001 |

### Determining the Multiplicity of Infection (MOI) based on Whole Genome Sequencing (WGS)

We first aimed to characterise the MOI using whole genome sequencing. A total of 230 RBC pellet samples at each timepoint were whole genome sequenced, resulting in 192 good-quality genomes (Figure 1 and Supplementary Table 1). To assess the MOI from these genomes, three algorithms were tested: *F_WS_*, THE REAL McCOIL and Runs of Homozygosity (RoH). All three algorithms showed robust correlation (Appendix 1). From here onwards any genome with *F_WS_*> 0.95 was considered monoclonal.

### Determining the MOI based on AmpSeq using gDNA from RBC pellets and plasma

We used the amplicon sequencing approach (AmpSeq) to assess MOI from paired RBC pellets and plasma samples obtained from the same individuals. The rationale was that the RBC pellet represents circulating parasites at a certain timepoint, while plasma could contain parasite DNA from previous schizont ruptures that is independent of sequestration. As described in the methods section, three markers were used (*cpmp*, *cpp*, *ama1*) and the marker showing the highest number of microhaplotypes in an individual was retained (*cpmp, cpp* and *ama1* in 34.8%, 34.8% and 30.4% of individuals, respectively, Supplementary Table 3). AmpSeq was successfully performed at each timepoint sample from 23 individuals with a polyclonal infection, as determined by WGS methods. 69 isolates were successfully genotyped in both RBC pellets and plasma gDNA, 7 were genotyped in RBC pellets gDNA only and 5 in plasma gDNA only (Supplementary Table 1).

In terms of sequence diversity, a total of 65 unique microhaplotypes were detected across all isolates. Among these, 98.5% (64) were identified in pellets, while 70.8% (46) were found in plasma. Furthermore, 19 microhaplotypes (29.2%) were exclusively present in pellets, while only one microhaplotype was only found in plasma. The mean MOI per isolate was significantly higher in RBC pellets compared to plasma (2.4 and 1.8 for pellets and plasma respectively, p = 0.0042) (Figure 2). Among isolates from the same individual, 66.7% (46/69) exhibited an identical number of genotypes detected when using gDNA from both RBC pellets and plasma. Furthermore, at parasitaemia over 1000 parasites/uL, the same MOI was detected in both plasma and pellet samples in 85.4% of the isolates (Supplementary Figure 1). This indicates the influence of parasite load on the sensitivity of AmpSeq when using plasma as the source of DNA. However, these data also show that plasma can be effectively used for parasite genotyping at relatively high parasitaemia levels.

**Figure 2.**
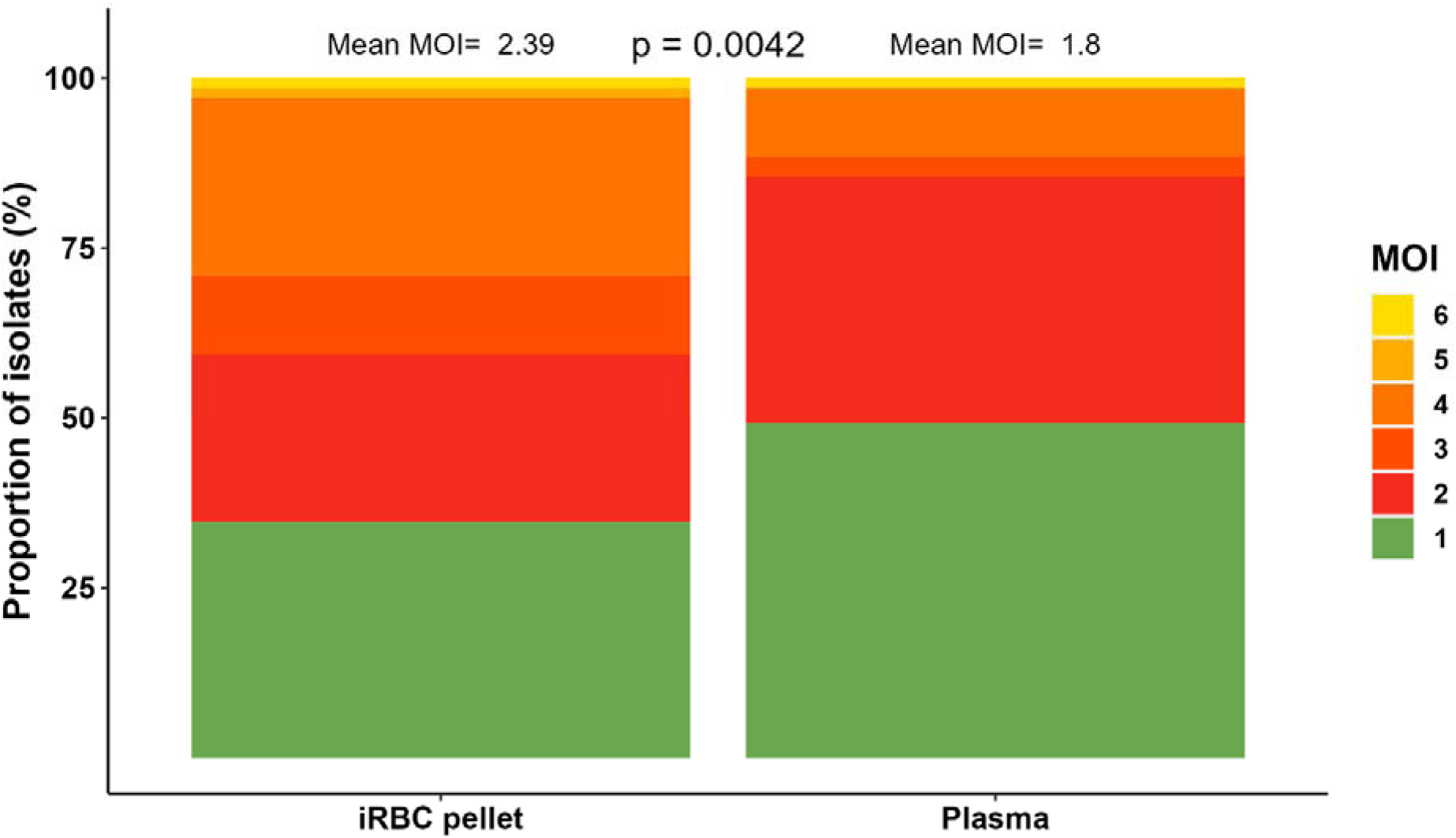
Comparison of the multiplicity of the infection determined using AmpSeq with gDNA from paired RBC pellet and plasma samples. The MOI per isolate, indicated as a colour scale, is the number of distinct microhaplotypes detected per isolate.

By visualizing the evolution of clonality at each time point (0h, 8h, 16h and 24h) during the 24h after treatment initiation (Figure 3), 7 infections showed the same MOI profile from RBC pellet and plasma over time (Figure 3A), while in 16 infections more microhaplotypes were detected in RBC pellet than in plasma (Figure 3B). Taken together, AmpSeq using gDNA from RBC pellets is more sensitive compared to AmpSeq using gDNA from plasma at low range parasitaemia. From here onwards, the AmpSeq MOI was defined as the number of microhaplotypes detected with gDNA from RBC pellets.

**Figure 3.**
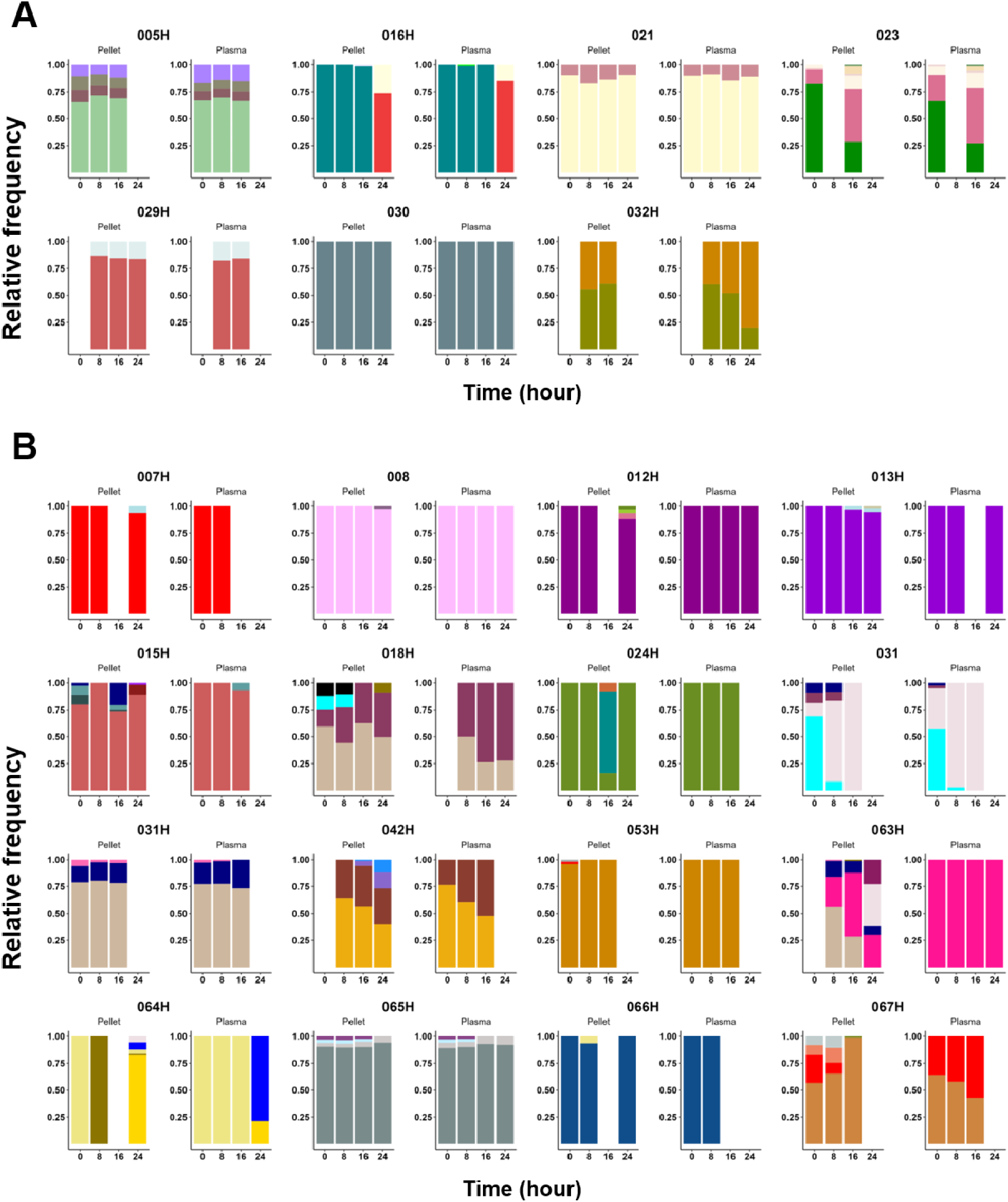
Relative proportion of microhaplotypes within patients, from AmpSeq using paired gDNA extracted from RBC pellets and plasma samples. A) Patients with the same microhaplotype profile between plasma and pellet gDNA. B) Patients with more microhaplotypes (higher MOI) detected within RBC pellet gDNA compared to plasma. Each colour indicates a unique microhaplotype within the whole population.

### Comparison of AmpSeq and WGS-based methods for determining MOI from pellet gDNA

A total of 68 isolates had MOI data available for both AmpSeq and WGS. We found a good correlation between AmpSeq and the different WGS-based methods (r= −0.61, r= 0.47 and r= −0.59 for *Fws*, THE REAL McCOIL and RoH, respectively) (Figure 4). In addition, 80.8% and 79.3% of the isolates showed concordant results (either monoclonal or polyclonal) between AmpSeq and *Fws*, or AmpSeq and RoH, respectively (Figure 4B). However, the AmpSeq method was able to detect up to 6 genotypes in an isolate, indicating greater sensitivity of AmpSeq in detecting multiple genomes in an isolate compared to WGS-based methods (Figure 4B). Furthermore, AmpSeq was the only method capable of quantifying the proportion of each genotype identified within an isolate. Together, these findings showed that the combination of different haplotyping techniques can be used to characterise genetic complexity quantitatively and qualitatively within an isolate. The MOI of a polyclonal isolate was then defined from the AmpSeq data.

**Figure 4.**
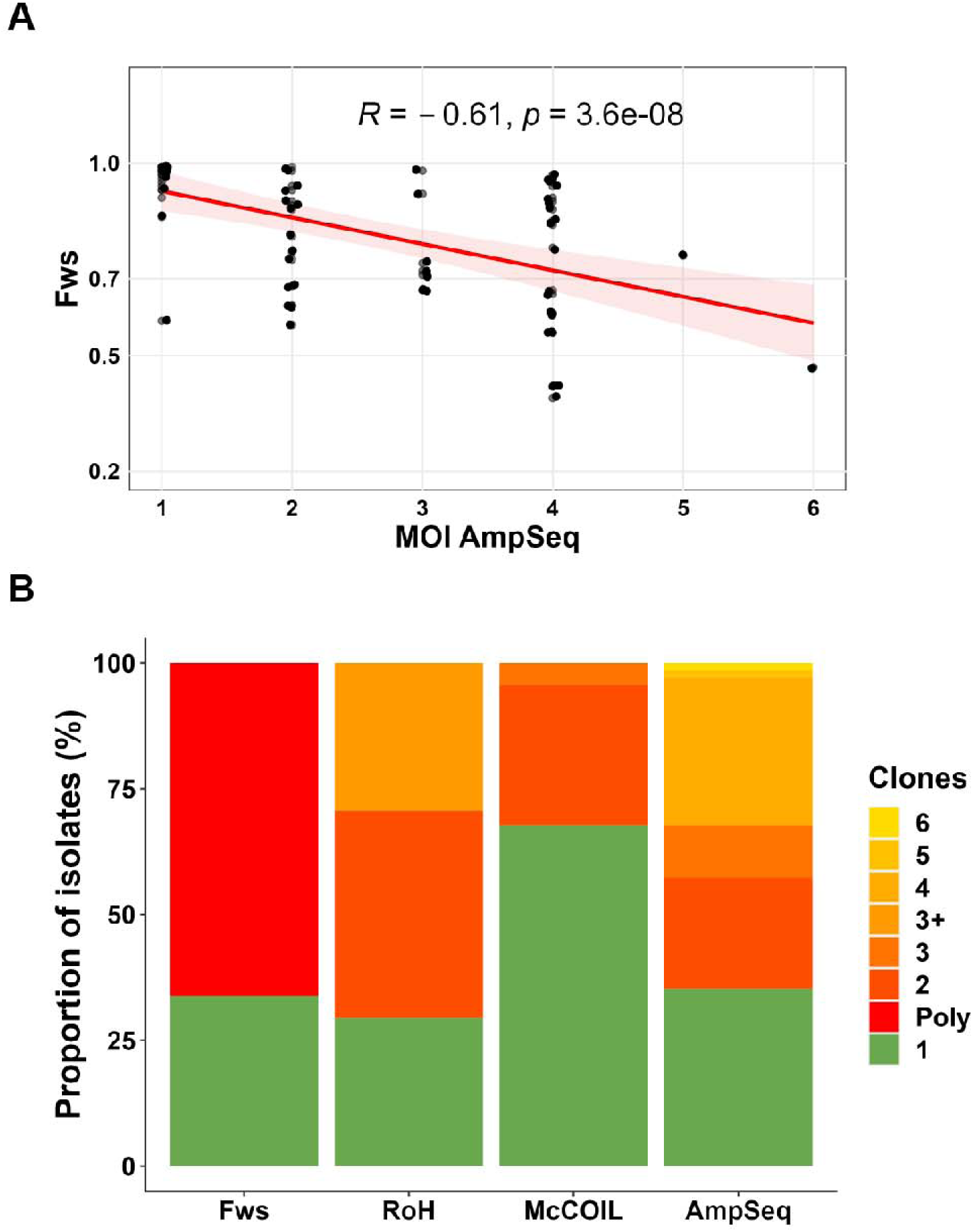
Comparison of AmpSeq to different WGS-based algorithms for determining the MOI. A) Correlation between *Fws* and AmpSeq. B) Proportion of isolates according to clonality in different genotyping techniques. Only individuals with at least one timepoint at *Fws* <0.95 were included in this analysis.

### Variability in parasite clearance rate among individuals and high rate of rebound in parasitaemia during antimalarial treatment

Having established the most sensitive method for MOI determination, we then aimed to characterise the clearance rate within the first 24 hours of treatment. Median parasite clearance time was faster in children treated with artesunate compared to those treated with quinine (Figure 5A), with the mean PC_50%_ of 8.3 and 12.2 hours, respectively (Figure 5B). There was a high variability in parasite clearance half-life within both treatment groups, ranging from 4.1 to 23.3 hours for artesunate and 4.0 to 24.0 hours for quinine (Figure 5B). Of the 48 individuals treated with artesunate, 95.6% (43/45) had a PC_50%_ below 16 hours, compared to 75% (12/16) of individuals treated with quinine (Figure 5B). The PC_50%_ could not be determined for four individuals (023, 014H, 029H, 063H) due to a non-significant decrease in parasitaemia (<50% reduction) between the first and last time points (Supplementary Data 2). Parasite clearance curves are within then same range as previous studies using quinine or artesunate [33] (Supplementary Data 2). These curves indicated substantial variation in initial clearance rates between individuals, highlighting the complex dynamics of parasite response to treatment in the early stage of infection management.

**Figure 5.**
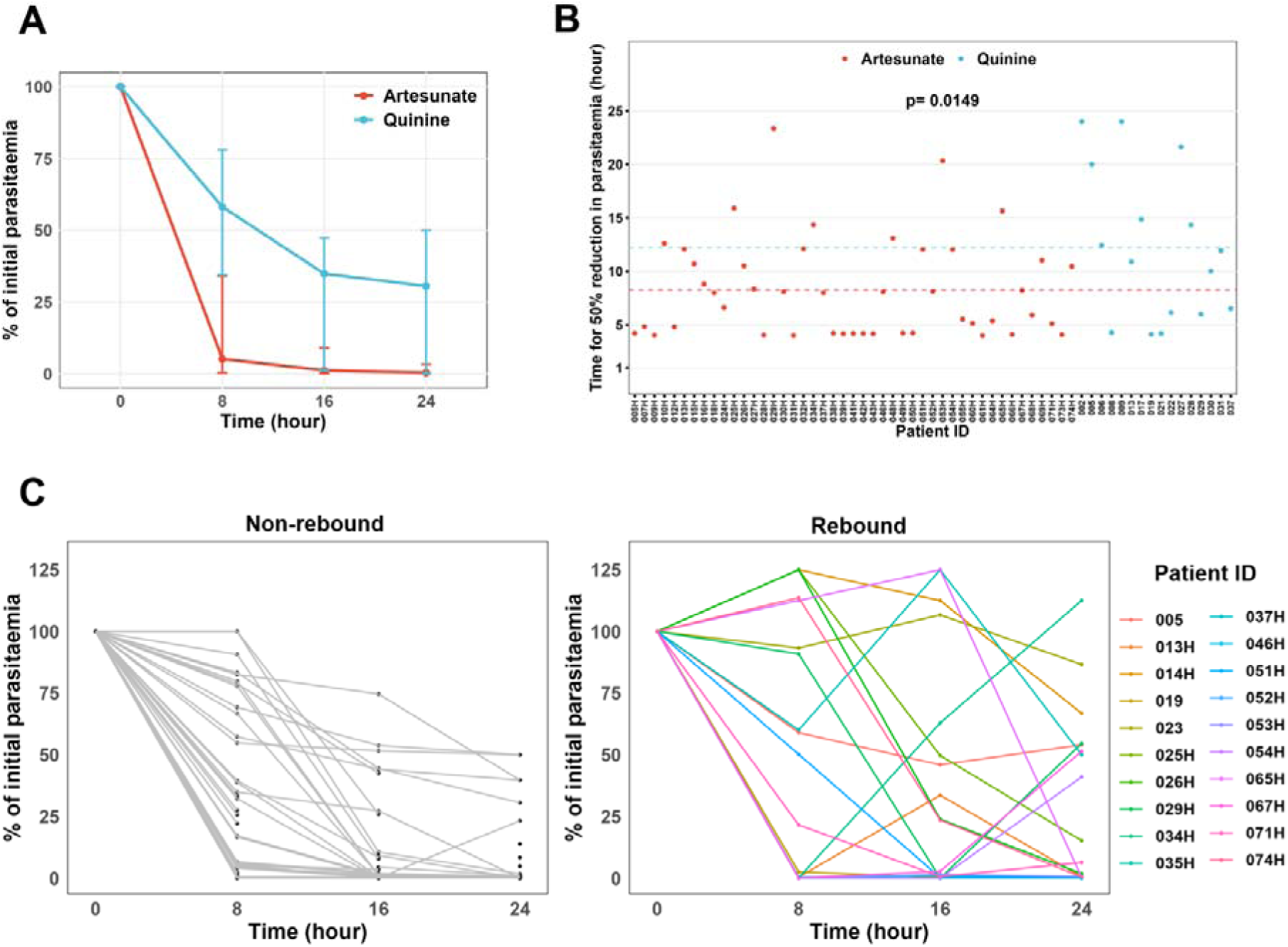
Parasite clearance curves of severe malaria cases treated with Artesunate or Quinine. A) Parasite clearance curve according to antimalarial treatment. B) Time for 50% reduction in parasitaemia according to treatment. C) parasite clearance curve highlighting the rebound in parasitaemia post-antimalarial treatment.

We took advantage of the time-series to investigate intra-host competition in polygenomic infections. Interestingly, we observed a ‘rebound’ in the parasitemia determined by microscopy reading. We defined rebound as an increase in parasite densities by at least 100 parasites/µL between two time points, and observed this phenomenon in 34.8% (20/65) of individuals (Figure 6C). Specifically, within the artesunate-treated group, 35.4% (17/48) of individuals experienced a rebound, while in the quinine-treated group, the rebound rate was 17.6% (3/17). These data indicate the potential release of sequestered parasites during anti-malarial treatment.

**Figure 6.**
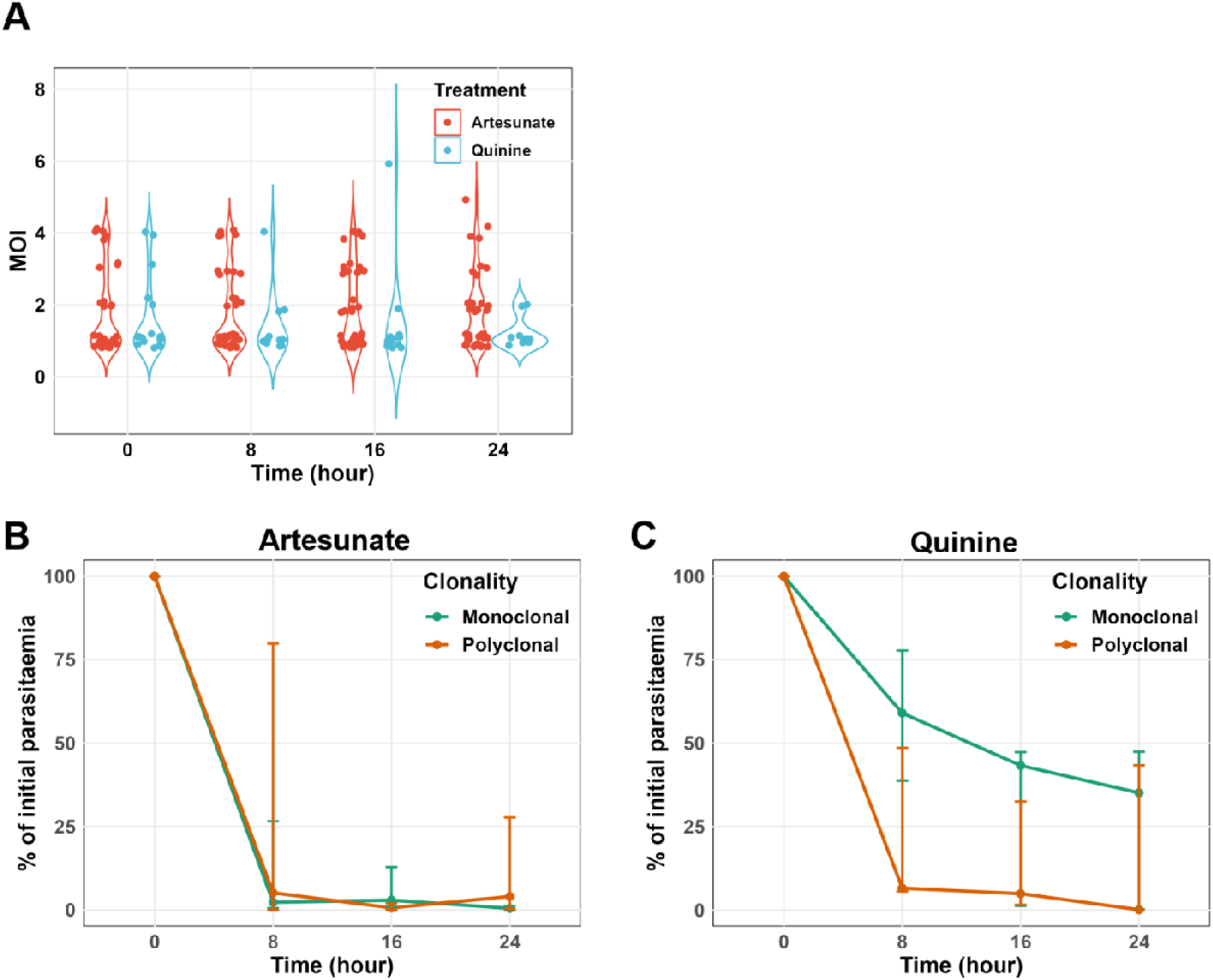
Dynamic of parasite clearance to parasite multiplicity of the infection. A) Dynamic of Multiplicity of Infection (MOI) determined by AmpSeq throughout the antimalarial treatment. Parasite clearance rate according to clonality at enrolment (H0) for artesunate (B) quinine (C) treatment groups.

### *P. falciparum* clearance rate is not related to MOI at the onset of the treatment

To determine whether the variability in the clearance rate of *P. falciparum* depends on the number of parasite genotypes in the infection, the dynamics of parasite clearance were compared between individuals with polyclonal and monoclonal infections during the first 24h of the severe malaria management under treatment pressure. Independently of the treatment administered, the mean MOI was relatively stable throughout the 24-hour period (Figure 6A). In individuals treated with artesunate, *P. falciparum* clearance showed no significant difference between polyclonal and monoclonal infections (Figure 6B). In contrast, although non-significant, treatment with quinine resulted in faster *P. falciparum* clearance in polyclonal infections (mean PC_50%_ of 4.3) compared to monoclonal infections (mean PC_50%_ of 12.4) (p= 0.1393) (Figure 6C). Part of this difference may be attributed to the higher median parasitaemia at enrolment in individuals treated with quinine and having monoclonal infections compared to those with polyclonal infections (2440000 vs 1520000 p/µL, p= 0.1924). In addition, there was no correlation between MOI at enrolment and parasite clearance rate (r= −0.06, p= 0.6494). These data show that the *P. falciparum* clearance rate does not depend on the initial parasite MOI.

### Sporadic detection of novel microhaplotypes during anti-malarial treatment

To measure the complexity of microhaplotypes within each isolate, we measured the Shannon entropy, a metric dependent on the number and proportion of each microhaplotype within an isolate. Despite the overall parasitaemia decreasing by an average of 335-fold over 24 hours, the microhaplotype entropy only slightly decreased between H0 and H8 (p= 0.0388), and remained stable afterwards (p= 0.6093) (Supplementary Figure 2). This stable MOI is explained by the detection of novel microhaplotypes that were not in circulation at H0. These ‘sporadic’ microhaplotypes were detected in over 60.9% (14/23) of participants, predominantly occurring from 16 hours after the start of treatment (Figure 7), likely indicating the release of sequestered parasites that were previously undetectable. Additionally, half of participants with sporadic microhaplotypes (007H, 008, 012H, 013H, 016H, 024h, 064H, and 066H) had monoclonal infections at H0 but displayed polyclonal infections at later time points (Figure 7A). However, most of the sporadic microhaplotypes were very minor genotypes (Figure 7A).

**Figure 7.**
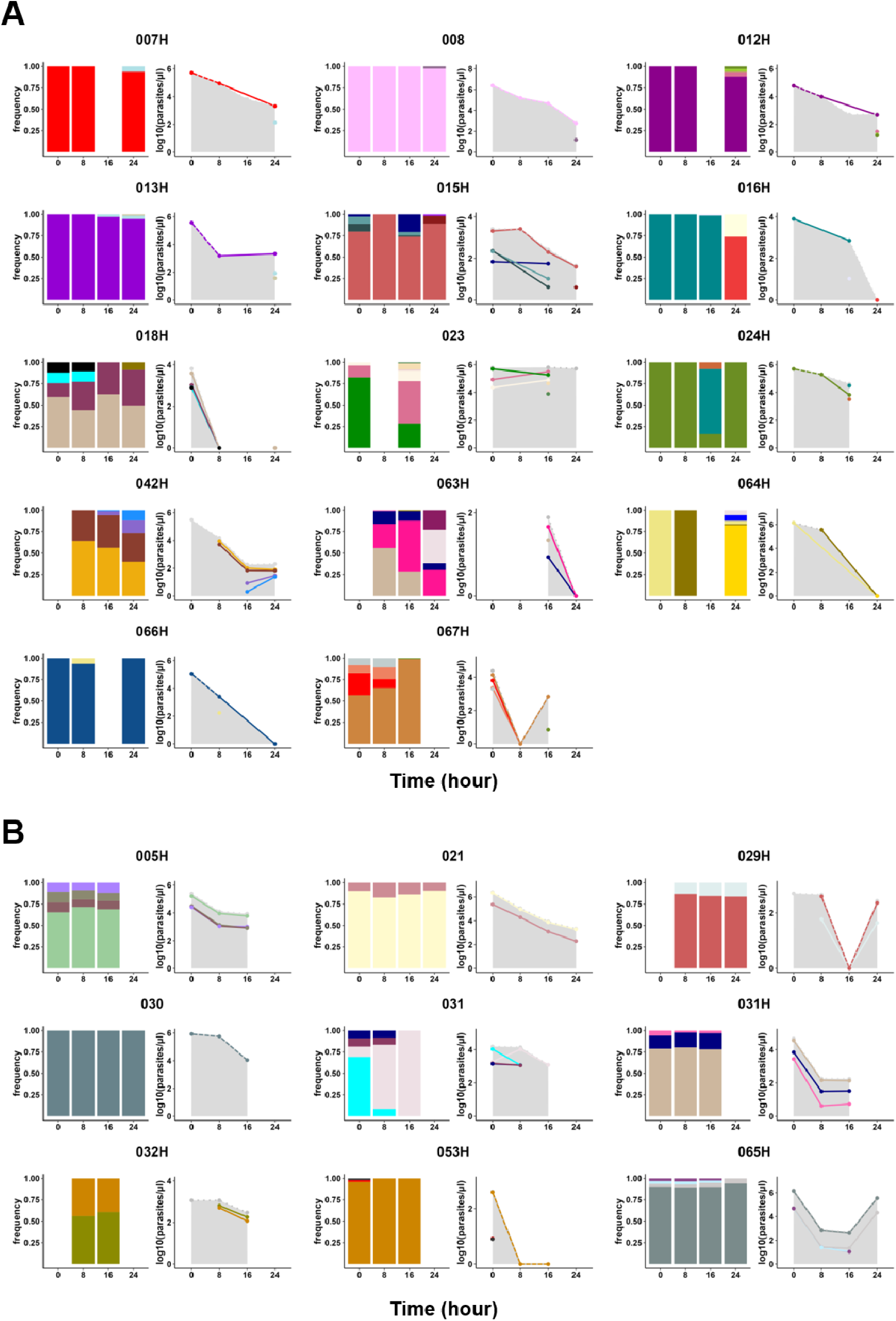
Subpopulation clearance curve during anti-malarial treatment, with (A) or without (B) sporadic detection of novel microhaplotypes. The left panel of each plot shows the relative abundance of microhaplotype subpopulations throughout the antimalarial treatment period. The right panel of each plot shows parasite densities of these subpopulations, calculated by multiplying the total parasitaemia by the relative abundance of the microhaplotypes. The clearance curves for the total parasite density are shown as a background surface (in grey). Each color represents a unique microhaplotype within the population. Sporadic detection is defined as the appearance of a new microhaplotype that was not present at H0.

We inferred the parasitaemia of each circulating genotype based on the total parasitaemia and the proportion of each microhaplotype. Notably, only one individual (067H) harbouring sporadic microhaplotypes showed a rebound in microhaplotype parasitaemia, indicating that the rebound in total parasitaemia observed was unrelated to the release of new genotypes in circulation after the start of the treatment.

Taken together, we here showed that, from blood samples taken on arrival at hospital (H0), it is usually not possible to genotype all *P. falciparum* microhaplotypes from an infection. About two third of the time, one or more distinct strain will appear within 24 hours, presumably as a result of release of sequestered *P. falciparum*.

### Parasite clearance is not correlated with drug resistance allele frequencies

We tested the hypothesis that drug sensitive genomes get cleared faster, by assessing allelic frequencies of resistance markers *crt*, *mdr1, kelch13, dhfr* and *dhps* over the 24 hours period. Of the 45 individuals for whom baseline drug resistance data was available, 95.6% (43/45) carried chloroquine-resistant allele (K76T) (Supplementary Figure 3A). However, the two individuals showing a mixture of sensitive and resistant alleles came from the group treated with artesunate (Supplementary Figure 3A). The frequency of the multidrug resistance gene 1 (*mdr1*) remained stable in the population throughout the 24 hours treatment period (Supplementary Figure 3B). Only one individual with a k13 mutation (V589I) at the time of enrolment was found in the study population (Supplementary Figure 3C). All isolates carried a triple mutation of the *dhfr* gene (N51I, C59R and S108N) (Supplementary Figure 3D). Similarly, 99% of isolates carried the A437G mutation in the *dhps* gene, with 23.2% and 0% a second mutation (Supplementary Figure 3E). Overall, we observed no alteration in genotypes across all analysed resistance markers within the same individual during antimalarial treatment.

## Discussion

In recent years, several approaches have been used to study the intra-host dynamics of polygenomic infections, including WGS which is the most comprehensive approach for genomic epidemiology, providing a complete picture of genetic variation. However, molecular markers, mainly AmpSeq, which combines several highly polymorphic antigen markers, is now the method of choice for high-resolution detection of MOI [22, 34]. We first aim in this study to evaluate different methods to study intra-host dynamics of polygenomic infections during a course of parasite clearance with an antimalarial treatment. Using the WGS data, we observed a robust correlation between the three algorithms (*Fws*, THE REAL McCOIL and RoH) used to determine the clonality of an isolate, in particular higher between *Fws* and RoH with 94.3% consistent results between the two metrics. However, 16.1% of polyclonal isolates according to *F_WS_* were classified as monoclonal according to THE REAL McCOIL. These data indicate that both the Fws and RoH algorithms exhibit greater sensitivity in detecting multiple infections compared to THE REAL McCOIL. Indeed, THE REAL McCOIL is a more conservative approach, which provides estimates that minimize overestimation of MOI by considering the likelihood of observed genetic variation being due to multiple parasite strains rather than sequencing errors or technical artefacts [24]. In this study, more than 79% of the isolates showed concordant results (either monoclonal or polyclonal) between AmpSeq and Fws or RoH. However, the AmpSeq method was able to detect up to 6 genotypes in an isolate, indicating greater sensitivity of AmpSeq in detecting multiple genomes in an isolate compared to WGS-based methods. Amplicon sequencing targeting highly polymorphic loci has demonstrated the capability to detect minority haplotypes with frequencies as low as 0.1% [26].

Previous studies have demonstrated that plasma can be used to detect and quantify the *Plasmodium* DNA by PCR [20, 21]. However, these studies also indicated that qPCR using whole blood DNA exhibits greater sensitivity than qPCR with plasma DNA. In line with the sequestration phenomenon of *P. falciparum* as shown by quantify the plasma levels of HRP2 or parasite DNA [18, 19], we hypothesized that genotyping parasites using DNA extracted from plasma might detect sequestered *P. falciparum* genotypes. This hypothesis assumes that plasma may contain parasite DNA released from prior schizont ruptures of all the parasite genotypes, which would be unaffected by sequestration at the time of blood sample collection. However, our findings revealed significantly higher microhaplotype complexity in the RBC pellet compared to plasma. These results indicate AmpSeq using gDNA from RBC pellets is more sensitive compared to AmpSeq using gDNA from plasma. To what extent the RBC pellet contains DNA from prior schizont ruptures remains to be determined. Consistent with previous findings where low levels of parasitemia was more effectively detected by qPCR using parasite DNA from whole blood compared to plasma [20], our study also found that parasite load influences the sensitivity of AmpSeq when using plasma as the DNA source. Indeed, the median parasitaemia of isolates with the higher MOI in the RBC pellet compared to plasma was 34.4 fold lower compared to the median parasitaemia of isolates with the same MOI in both biological materials. Interestingly, at parasitaemia levels exceeding 1000 parasites/µL, the same multiplicity of infection (MOI) was detected in both plasma and pellet samples in 85.4% of the isolates. This indicates that plasma can be effectively used for parasite genotyping in retrospective studies.

We observed high variability in parasite clearance half-life within both treatment groups with faster parasite clearance rate in children treated with artesunate compared to those treated with quinine. Artesunate is a fast-acting antimalarial drug with a pronounced effect on ring-stage parasites, whereas quinine primarily targets mature trophozoites [35]. Rapid clearance results from artesunate action on the circulating ring-stage parasites and their subsequent removal predominantly by the spleen, preventing the parasite sequestration [36, 37]. In line with our findings, a recent study of Ghanaian children showed a prolonged delay in parasite clearance among children with severe malaria, even after three days of treatment with artesunate [38]. This increased time to parasite clearance was associated with age, low haemoglobin levels and high number of previous malaria diagnoses [38]. Other studies have noted variations in parasite clearance rates in children treated for uncomplicated falciparum malaria with artemisinin-based combination therapy, both in Kenya and Tanzania [39, 40]. These findings suggest that factors other than drug resistance may contribute to slower parasite clearance in certain infections, as there is no evidence of artemisinin resistance in these regions. It has been shown that, in areas where artemisinin resistance is not present, the parasite clearance time can be affected by factors such as the age of the host, host acquired immunity, initial parasitemia levels, and the developmental stages of parasite [11–13]. Furthermore, although controversial, studies have shown a link between the genetic complexity of *P. falciparum* and delayed parasite clearance in Africa [14, 15]. Our data show that the *P. falciparum* clearance rate in children with severe malaria independent of parasite MOI at the onset of treatment. Notably, *P. falciparum* clearance was faster in polyclonal infections in quinine-treated children. However, this difference was attributed to significantly higher parasitemia at enrolment in individuals treated with quinine.

Interestingly, during antimalarial treatment, a high rate of “rebound” in total parasitemia and specific parasite subpopulations parasitemia was observed. Specifically, an increase in total parasitemia between two time points was detected in 34.8% of individuals. In four individuals (023, 031, 063H, and 064H), minor genotypes present at baseline (H0) increased in relative frequency, becoming the dominant microhaplotype. Additionally, sporadic detection of microhaplotypes occurred in over 60.9% of participants, predominantly from 16 hours after treatment initiation. Half of the participants with sporadic microhaplotypes had monoclonal infections at baseline but exhibited polyclonal infections at subsequent time points. These findings potentially highlight release of sequestered *P. falciparum* genotypes during antimalarial treatment [34] Interestingly, none of these sporadic microhaplotypes were detected in pellet or plasma samples at H0, despite the much higher parasite density at that timepoint that would have facilitated detection of a rare genotype. Either these microhaplotypes recently emerged from the liver, with no or little parasite DNA in the plasma yet, or, probably more likely, these sporadic microhaplotypes are present at extremely low parasitaemia making them undetectable in plasma, and were all sequestered at H0.

Additionally, a study by Marks et al. (2005) reported a parasitological rebound effect in infants treated with a single dose of sulfadoxine-pyrimethamine, attributed to the selection of drug-resistant parasites shortly after drug clearance [41]. However, no mutation associated with artemisinin resistance has yet been identified in Benin [9, 10]. In addition, no alterations were observed in allele frequencies across all analysed resistance markers within the same individual post-antimalarial treatment. Two other studies using AmpSeq detected sporadic microhaplotypes or an increase in the frequency of minor microhaplotypes during the treatment of uncomplicated *P. falciparum* infections in regions with no signs of artemisinin resistance [39, 40]. In *falciparum* malaria, parasitized red blood cells circulate in the peripheral blood for only one-third of the 48-hour asexual cycle [42]; for the remainder of the cycle, they are sequestered in the venules and capillaries. For these infections, the peripheral blood parasitemia at a given time point does not reflect the total parasite burden.

Limitations of this study include the lack of sampling timepoints beyond 24 hours post-treatment, which affected the accurate determination of parasite clearance half-life for some individuals using the standard WWARN method [43]. This was due to the fact that the parasitemia reduction rate was less than 50% of the initial parasitemia at the last recorded timepoint. Therefore, we used linear interpolation to estimate the time required for a 50% reduction in parasitaemia, referred as the extrapolated parasite clearance half-life (PC_50%_). However, unlike therapeutic efficacy studies typically focusing on day 3, 7 and 21/28, we were able to monitor the dynamics of *P. falciparum* complexity during the initial management of severe malaria.

This study demonstrates that combining different haplotyping techniques can effectively determine genetic complexity. However, AmpSeq is the most sensitive approach and should be the first choice for genotyping to monitor the evolution of genotype clearance following antimalarial treatment. Additionally, plasma can be effectively used for parasite genotyping at sufficient parasitaemia levels. We demonstrated that genotyping a blood sample from a patient on arrival at hospital is typically not sufficient to detect all parasites genotypes present in the infection.

## Conflicts of interest

No conflicts of interest were reported by the authors.

## Financial support

This work was funded by grants from the French National Research Agency (18lJCE15lJ0009lJ01), the ATIP-Avenir programme and by Institut Merieux. The “SPFlJPost doc en France” programme of the “Fondation pour la Recherche Médicale” funded the author “Balotin Fogang” (SPF202209015889).

## Ethical approval

Ethical clearance was obtained from “Comité d’Ethique de la Recherche CER_ISBA Benin” (clearance n°90, 06/06/2016 and clearance n°38, 16/05/2014).

## Data Availability

All data produced in the present work are contained in the manuscript or are available online at the European Nucleotide Archive (ENA) under the study accession number PRJEB2136 and on Zenodo at https://zenodo.org/records/13224728.

https://www.ebi.ac.uk/ena/browser/view/PRJEB2136

https://zenodo.org/records/13224728

## Acknowledgments

We would like to thank the participants and their parents/guardians for granting approval to participate in this study. We thank the staff of the paediatric wards at the CHUMEL hospital in Cotonou and at Saint Joseph medical centre in Sô-Tchanhoué, Benin, for their help. Our thanks go to MalariaGen for the whole genome sequencing and to the GenSeq platform for the amplicon sequencing.

## Author contributions

Conceptualization by A.C and GI.B. Field work and sample processing by C.K, S.E and G.A. Data analysis by B.F, E.G. Writing of original draft by B.F, E.G. Reviewing and editing of manuscript by B.F., E.G, C.K, GI.B, A.C. Funding acquisition by A.C and GI. B.

## Data availability

All whole-genome sequencing (WGS) data generated and analyzed in this study have been deposited in the European Nucleotide Archive (ENA) under the study accession number PRJEB2136, with individual accession numbers in Supplementary Table 1. The raw reads from the Amplicon sequencing (AmpSeq) data are publicly available on Zenodo (https://zenodo.org/records/13224728), with corresponding individual accession numbers provided in Supplementary Table 3. The raw data and complete AmpSeq results are included as Supplementary Tables 1 and 2, respectively.

## Description of supplementary Material

**Supplementary Figure 1. Parasitaemia according to the concordance between MOI from paired RBC pellet and plasma samples, determined by AmpSeq.** “Pellet-plasma match” are isolates with identical MOI in plasma and pellet samples. “Low in plasma” are isolates in which the measured MOI was lower in plasma compared to pellet samples. There were only 2 isolates with MOI higher in the plasma than in the pellet, which are not shown on this figure.

**Supplementary Figure 2. Microhaplotype entropy over time during antimalarial treatment.**

**Supplementary Figure 3. Frequency of antimalarial drug resistance markers throughout treatment.**

**Supplementary Table 1. Epidemiological data of study participants**.

**Supplementary Table 2. PCR primers sequences and amplification conditions.** List of forward and reverse primers used for AmpSeq and the amplification program.

**Supplementary Table 3. Amplicon Sequencing results.** AmpSeq read counts and frequencies, for three markers (*cpmp*, *ama1-D3*, *cpp*) used in both plasma and red blood cell pellets. The table identifies the selected marker for each individual, with the marker displaying the highest number of microhaplotypes being retained. The last column indicates ‘sporadic’ microhaplotypes, defined as novel microhaplotypes not detected at H0.

**Appendix 1. Determining the Multiplicity of Infection (MOI) based on Whole Genome Sequencing (WGS).**

**Supplementary Data 1. Non-reference allele frequency (NRAF) distribution across all heterozygous SNPs (left) and heterozygosity calculated in 100-kb bins (right) to highlight RoH.**

**Supplementary Data 2. Individuals parasite clearance curve by timepoints post-antimalarial treatment. Quinine (A) and artesunate (B) treated groups.**

## Notes

### Competing Interest Statement

The authors have declared no competing interest.

### Funding Statement

This work was funded by grants from the French National Research Agency (18‐CE15‐0009‐01), the ATIP-Avenir programme and by Institut Merieux. The SPF‐Post doc en France programme of the Fondation pour la Recherche Médicale funded the author Balotin Fogang (SPF202209015889).

### Author Declarations

Ethics committee/IRB of "Institut des Sciences Biomedicales Appliquees Benin" (clearance number 90, 06/06/2016 and clearance number 38, 16/05/2014) gave ethical approval for this work.

