## Supplementary Material for "Detection of novel *Plasmodium falciparum* haplotypes under treatment pressure in pediatric severe malaria"

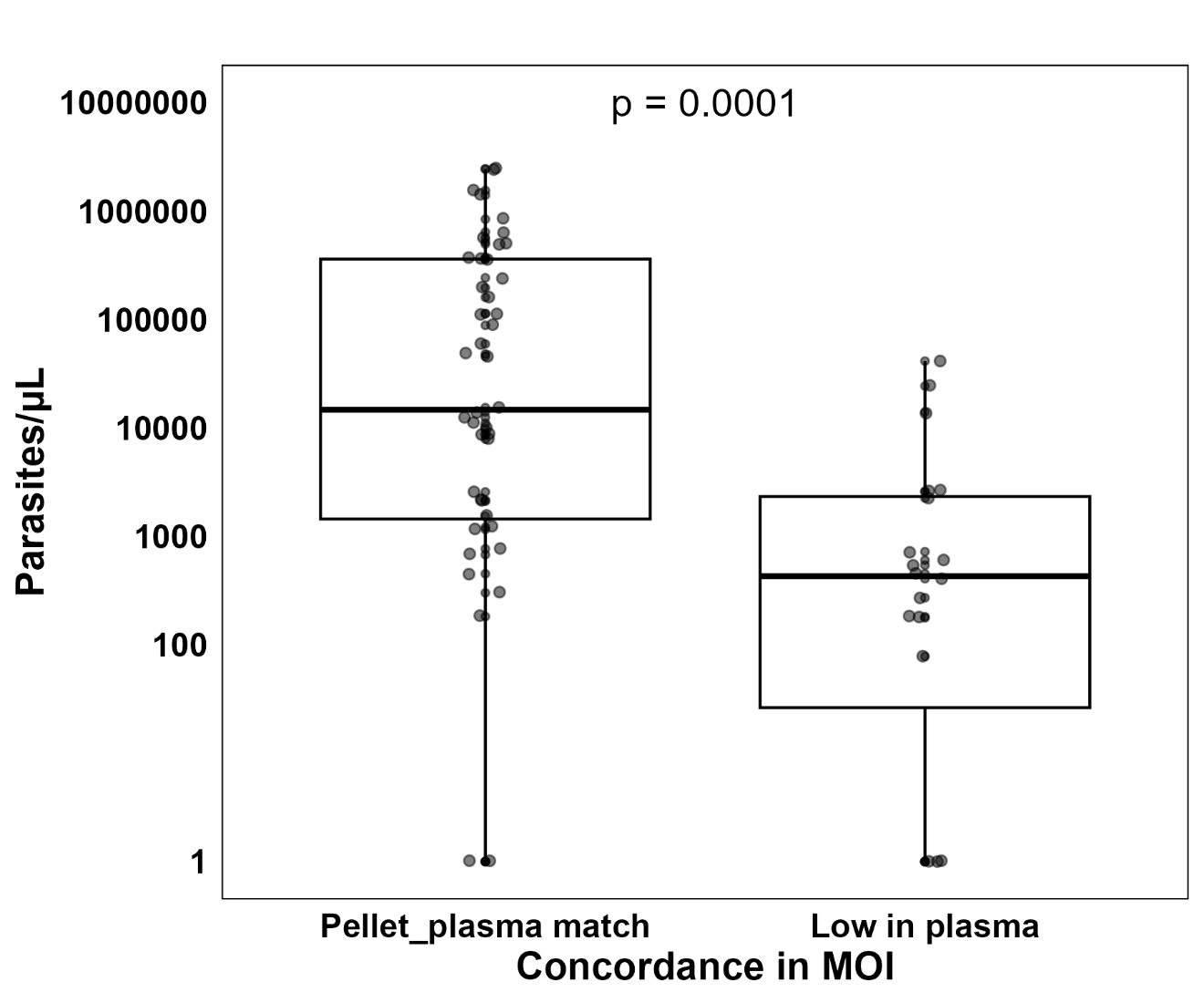

**Supplementary Figure 1. Parasitaemia according to the concordance between MOI from paired RBC pellet and plasma samples, determined by AmpSeq.**

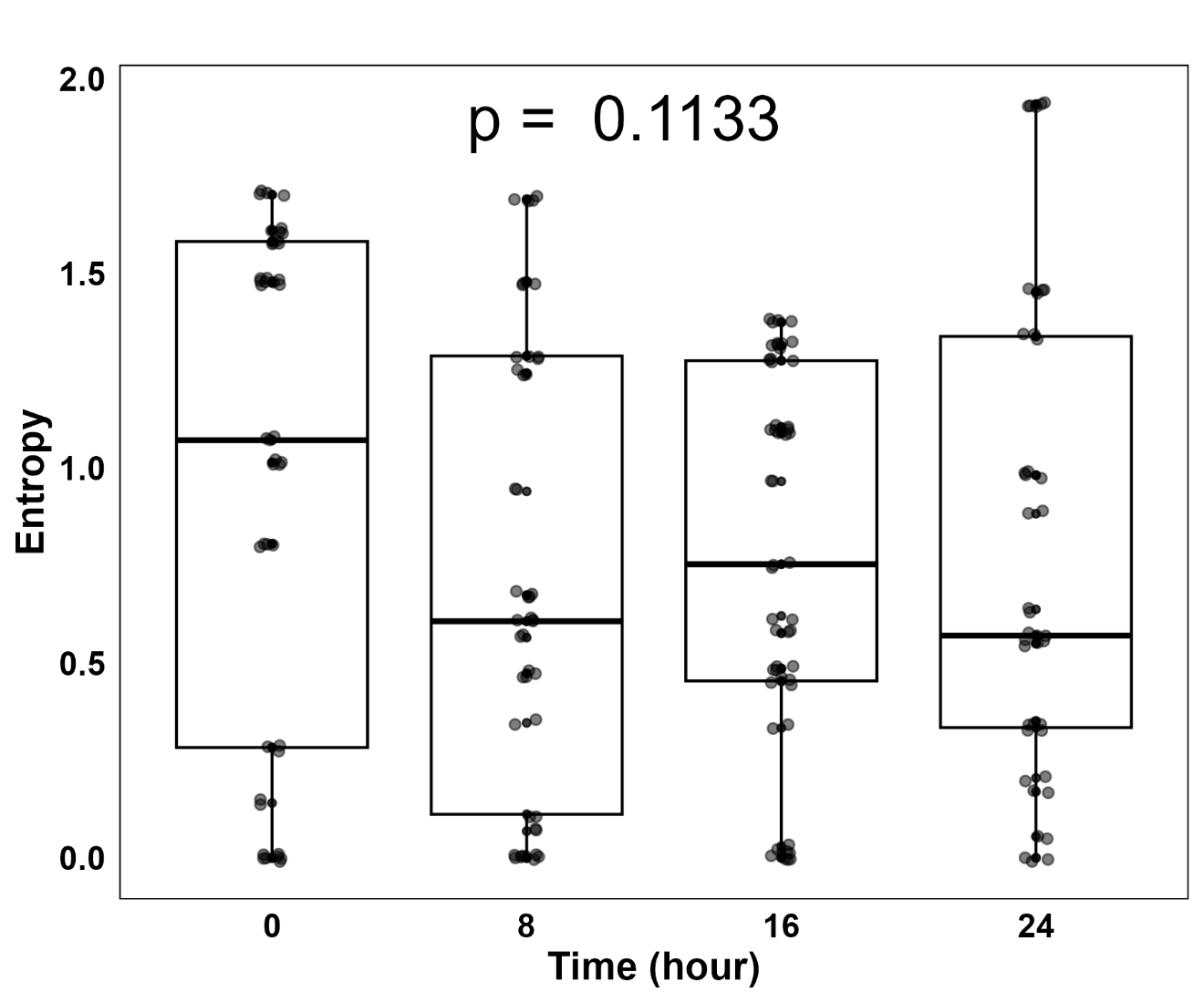

**Supplementary Figure 2. Microhaplotype entropy over time during antimalarial treatment.**

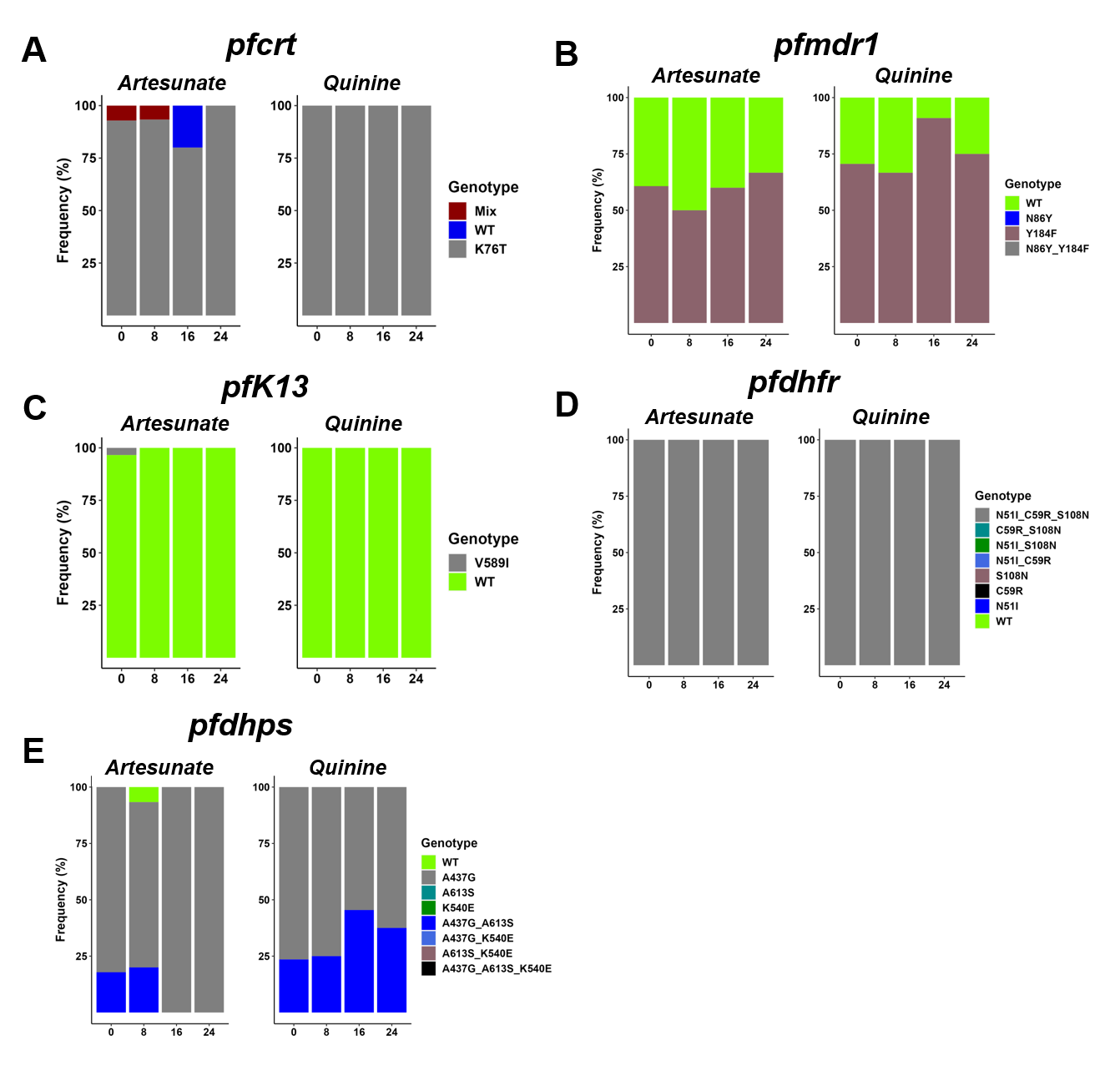

**Supplementary Figure 3. Frequency of antimalarial drug resistance markers throughout treatment.**

**Supplementary Table 2. PCR primers sequences and amplification conditions.**

| **Primers** | **Amplification Conditions** |
| --- | --- |
| **Primary PCR** | **Primary PCR** |
| cpmp_prim_fw: CGATACAGGACATATAGA  cpmp_prim_rv: TTCAATAACATTTACTAGG  cpp_prim_fw: TGTCTGAACCAAATTCAA  cpp_prim_rv: GAATTTGTCACATTTGATGA  ama1-D3_prim_fw : GTTTAATTAACAATTCATCATAC  ama1-D3_prim_rv: GTGTTGTATGTGATGCTC | 95°C for 3′, 20 cycles of 98°C for 20″ and 54°C for 15″ and 72°C for 45″, followed by 72°C for 2′ |
| **Nested PCR** | **Nested PCR** |
| cpmp_nested: CATAAGTCATTAAAATTTATGGAT  cpmp_nested: CGTTACTATCAAGATCGTTAATAT  cpp_nested: CAAGTTCACTTTTGGGAAATG  cpp_nested: ATTACTACCTTTCAGCATATCCGA  ama1-D3_ nested: TACTACTGCTTTGTCCCATC  ama1-D3_ nested: TCAGGATCTAACATTTCATC | 95°C for 3′, 10 cycles of 98°C for 20″ and 55 °C for 15″ and 72 °C for 45″, 10 cycles of 98 °C for 20″ and 68°C for 15″ and 72 °C for 45″followed by 72 °C for 1′30″ |

fw: Forward. rv: Reverse.

**Appendix 1: Determining the Multiplicity of Infection (MOI) based on Whole Genome Sequencing (WGS)**

To assess the MOI from the 192 good-quality genomes genomes, three algorithms were tested: *F_WS_*, THE REAL McCOIL and Runs of Homozygosity (RoH). Any genome with *F_WS_* > 0.95 was considered monoclonal. The RoH method categorises isolates as monoclonal (one dominant genotype), biclonal (2 dominant genotypes) or multiclonal (3+ dominant genotypes) (Figure B). THE REAL McCOIL method estimates the actual number of genotypes present in an isolate. We observed a robust correlation between the three methods, particularly higher *Fws* and RoH (r= 0.78, p < 0.0001) (Figure A). In addition, 94.3% (181/192) of isolates showed consistent results (either monoclonal or polyclonal) between *Fws* and RoH (Figure C). However, 83.9% (161/192) of the isolates showed consistent results (either monoclonal or polyclonal) for both *Fws* and THE REAL McCOIL, while 16.1% (31/192) were polyclonal according to *F_WS_* but monoclonal according to THE REAL McCOIL, making the latter algorithm more conservative (Figure C). These data indicate that both the *Fws* and RoH algorithms exhibit greater sensitivity in detecting multiple infections compared to THE REAL McCOIL. By WGS-based methods, 52.3% (34/65) of the infections were polyclonal for at least one timepoint over the 24 hours follow-up.

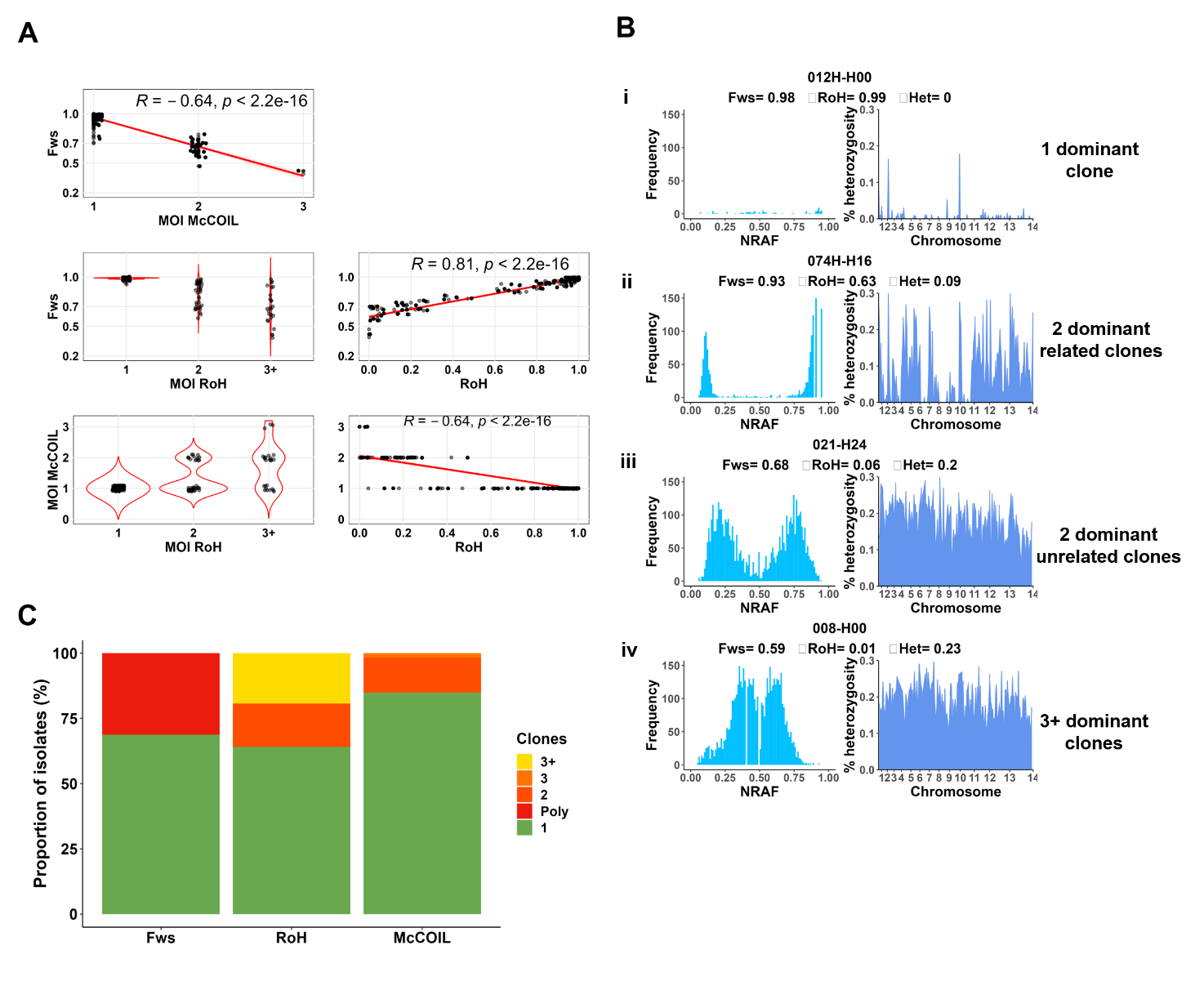

**Figure.** **Comparison of different approaches to determine the multiplicity of the infection**. A) Correlation between three different WGS-based algorithms (Fws, McCOIL, RoH) used to determine the MOI from 192 genomes. B) Four illustrative samples showing the non-reference allele frequency (NRAF) distribution across all heterozygous SNPs (left) and heterozygosity calculated in 100-kb bins (right) to highlight RoH. Sample i is clonal, with within-sample heterozygosity (Het) of 0 and Fws at 0.98. Sample ii and iii are likely two genotypes, as evidenced by the bimodal distribution of NRAF. However only sample ii is showing runs of homozygosity (RoH), indicating that the 2 genotypes within that sample are genetically related. Sample iv contains a complete mixture of genotypes, at least partly unrelated. All genomes are available in Supplementary Data 1. C) Clonality levels according to WGS-based algorithms.

**Supplementary Data 1. Non-reference allele frequency (NRAF) distribution across all heterozygous SNPs (left) and heterozygosity calculated in 100-kb bins (right) to highlight RoH.**

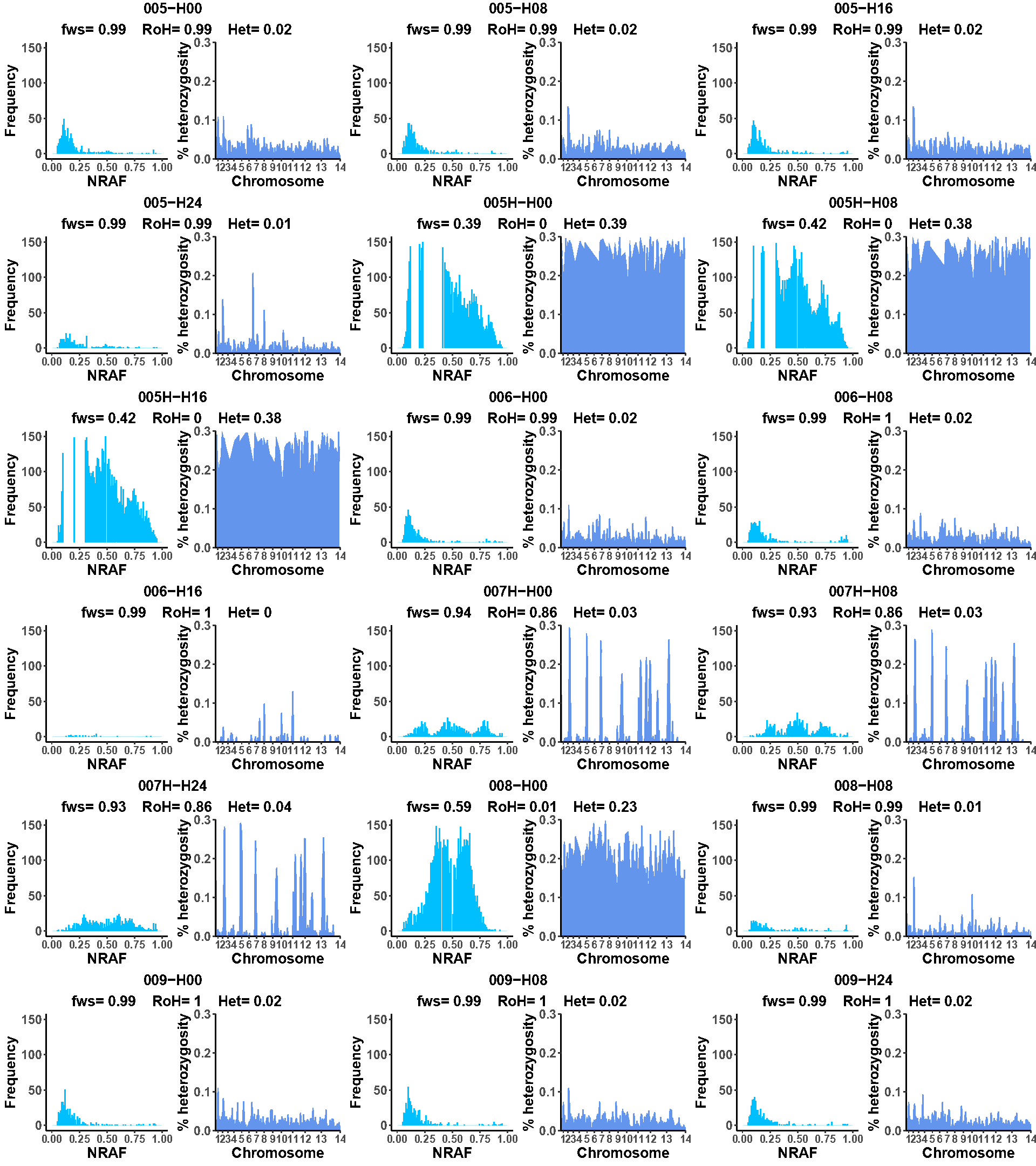

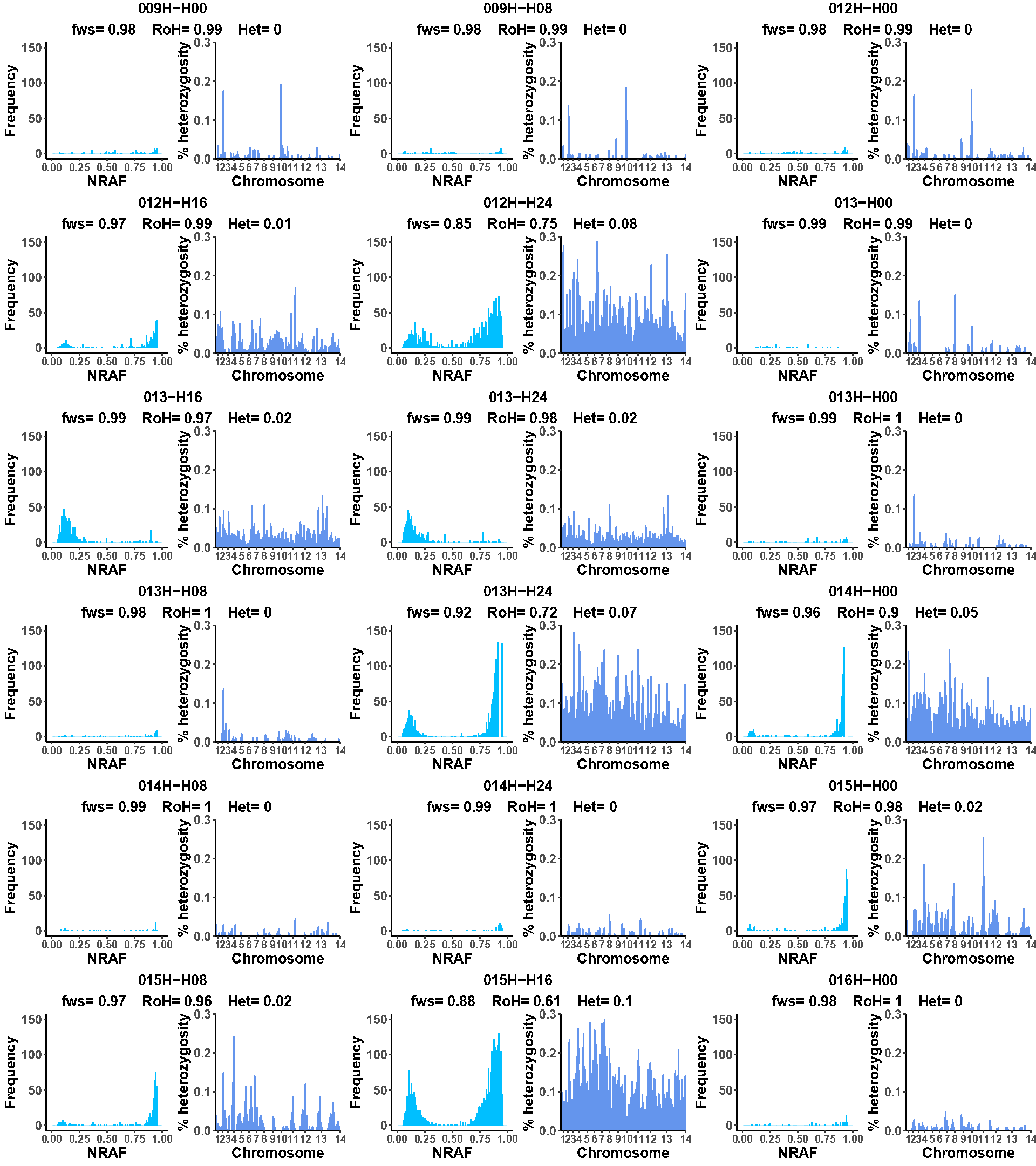

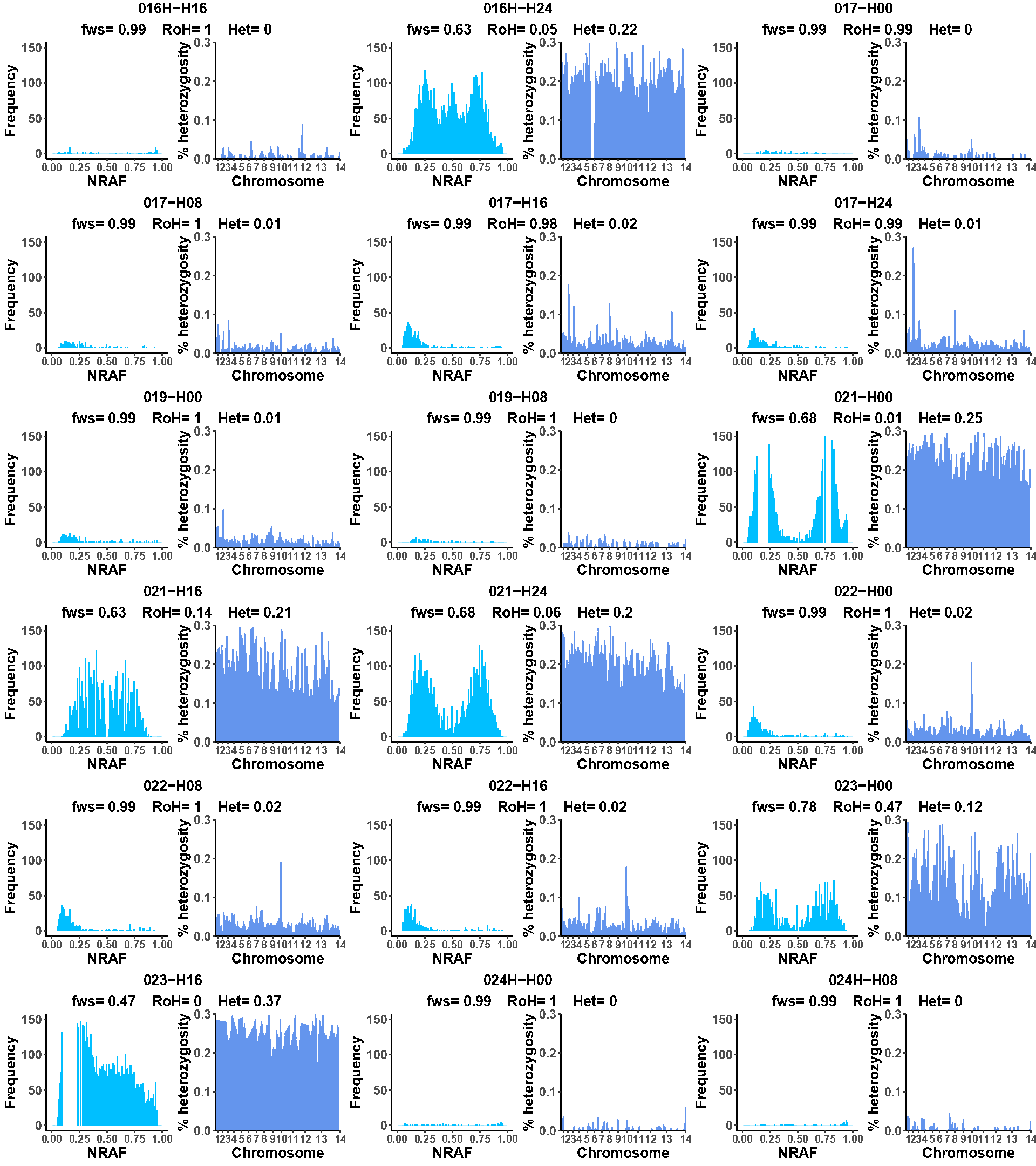

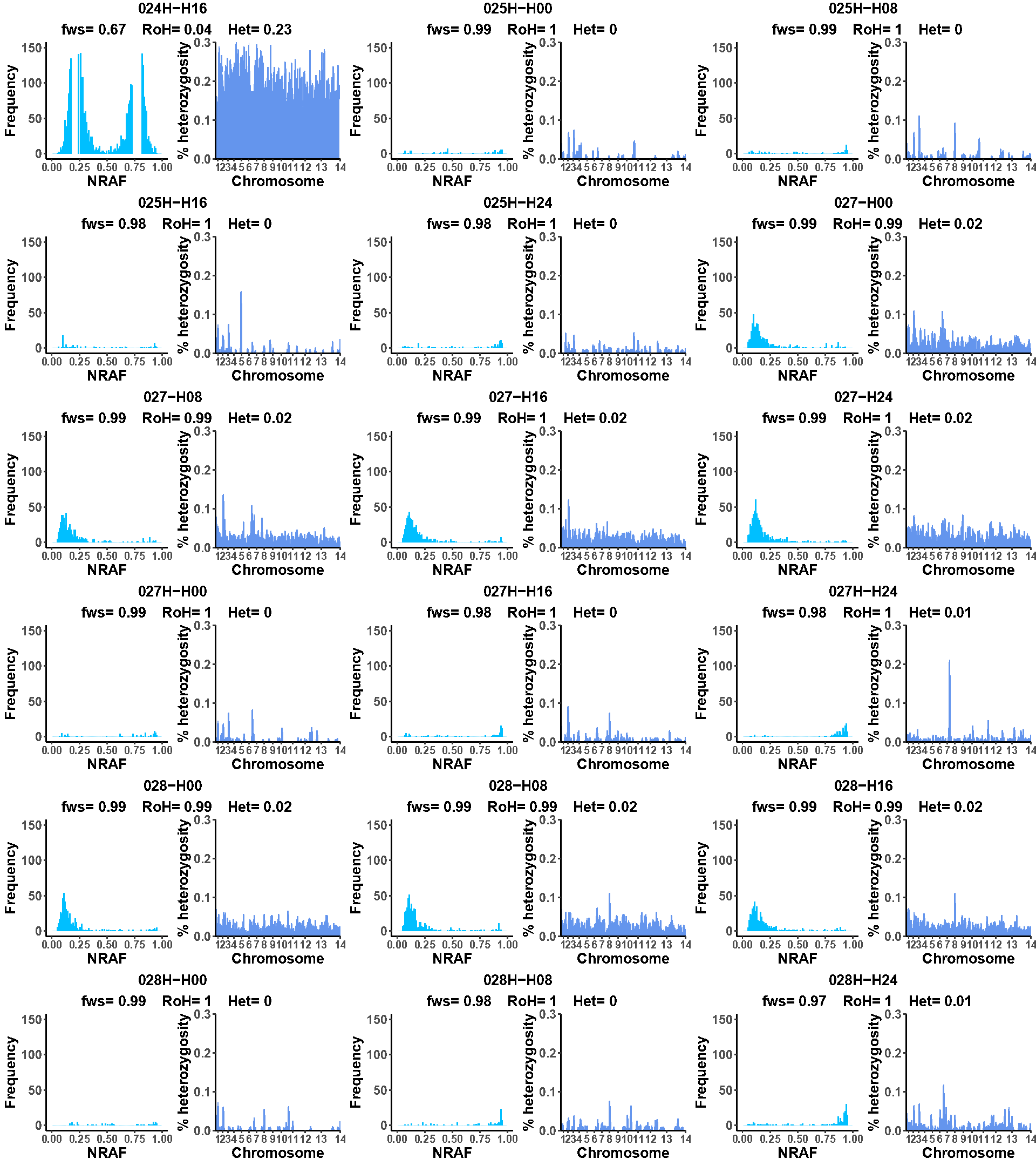

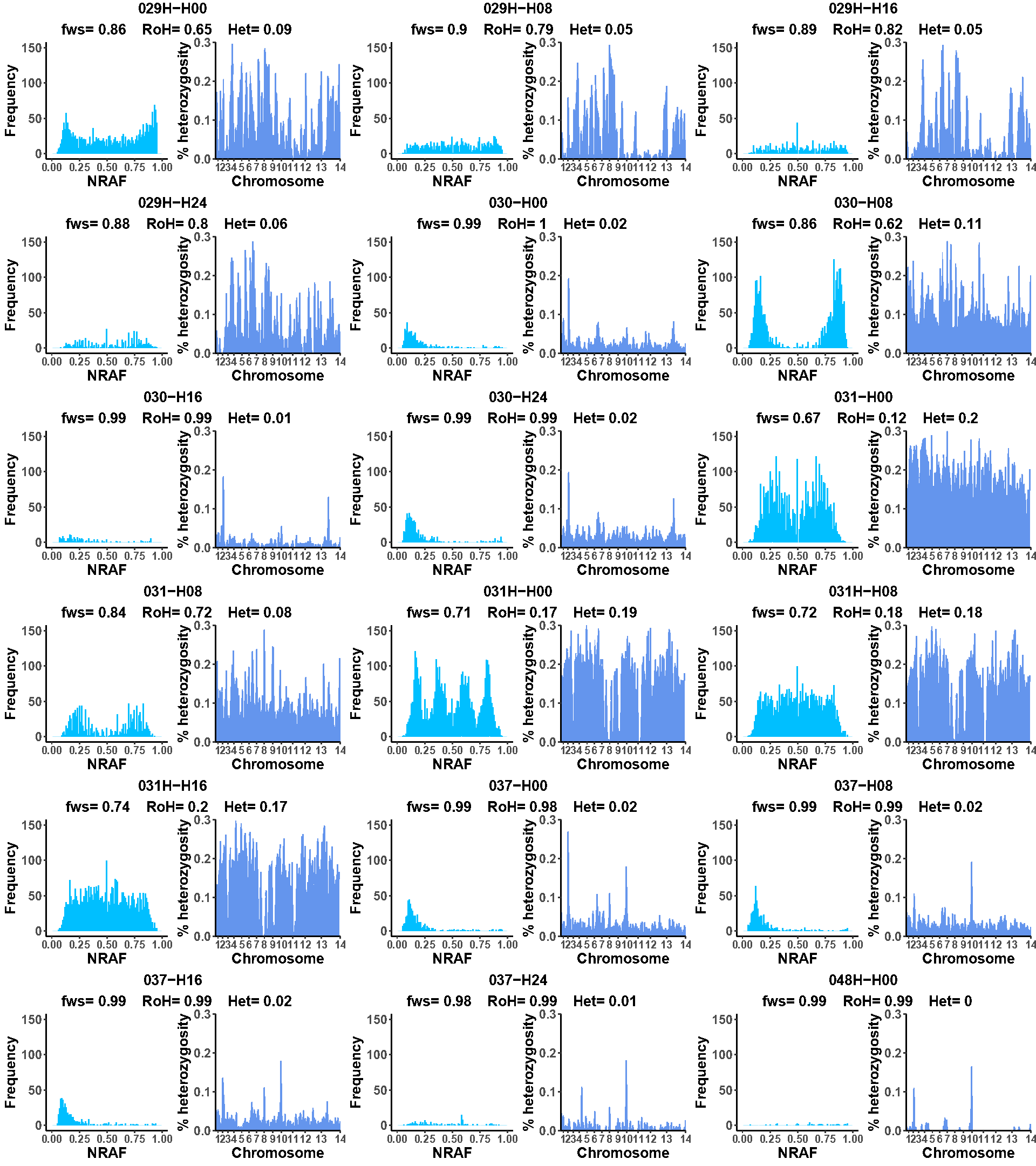

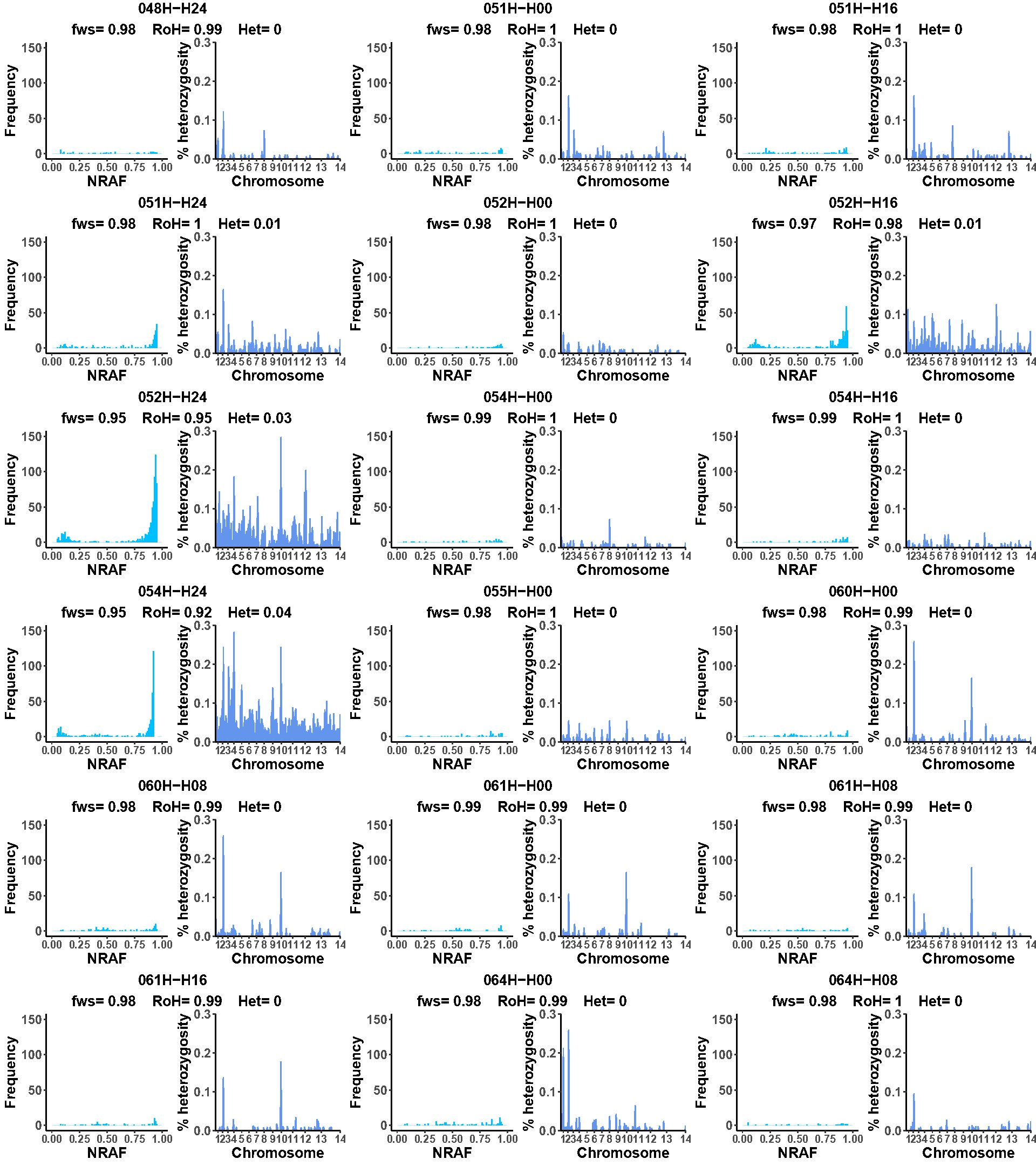

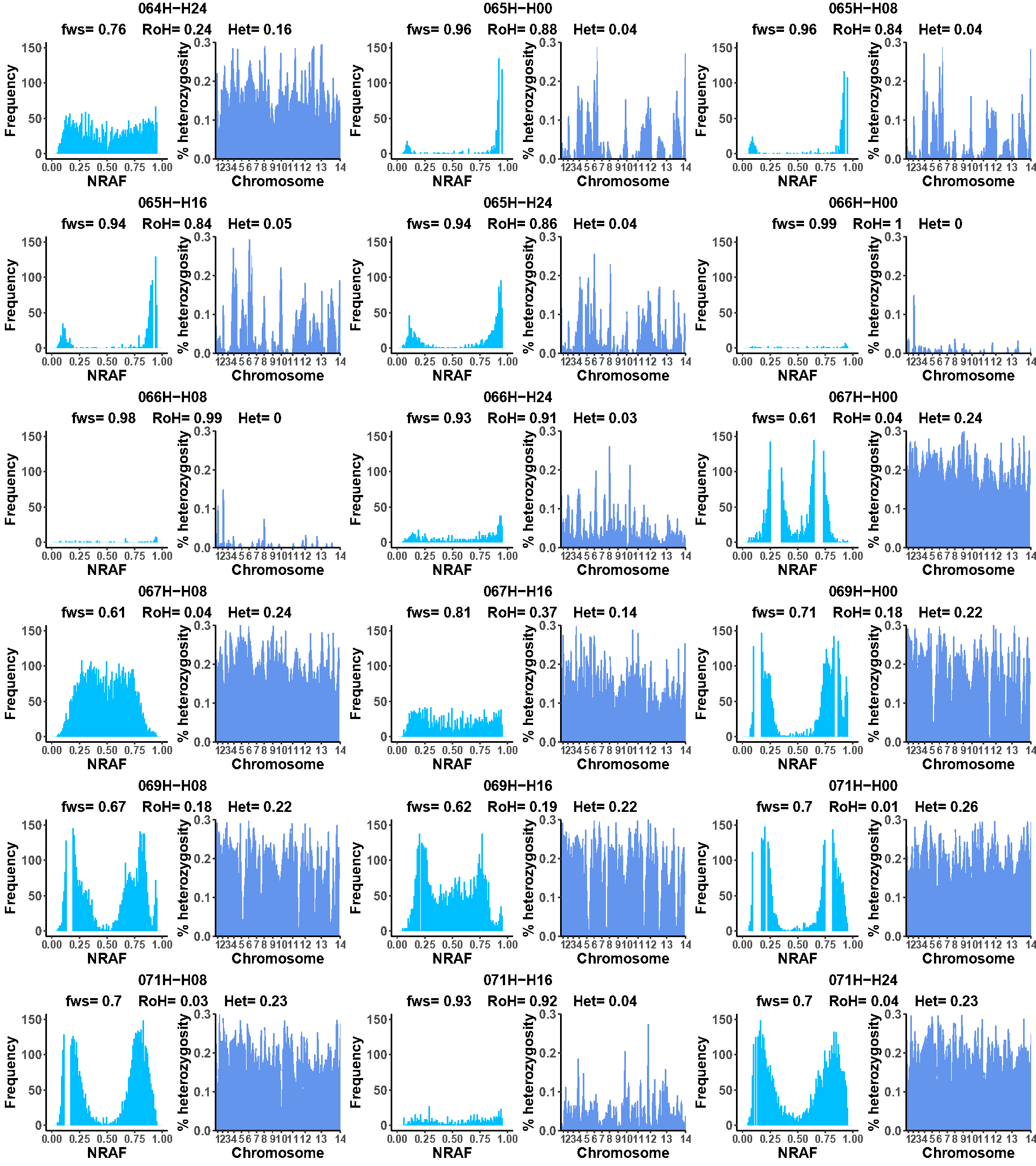

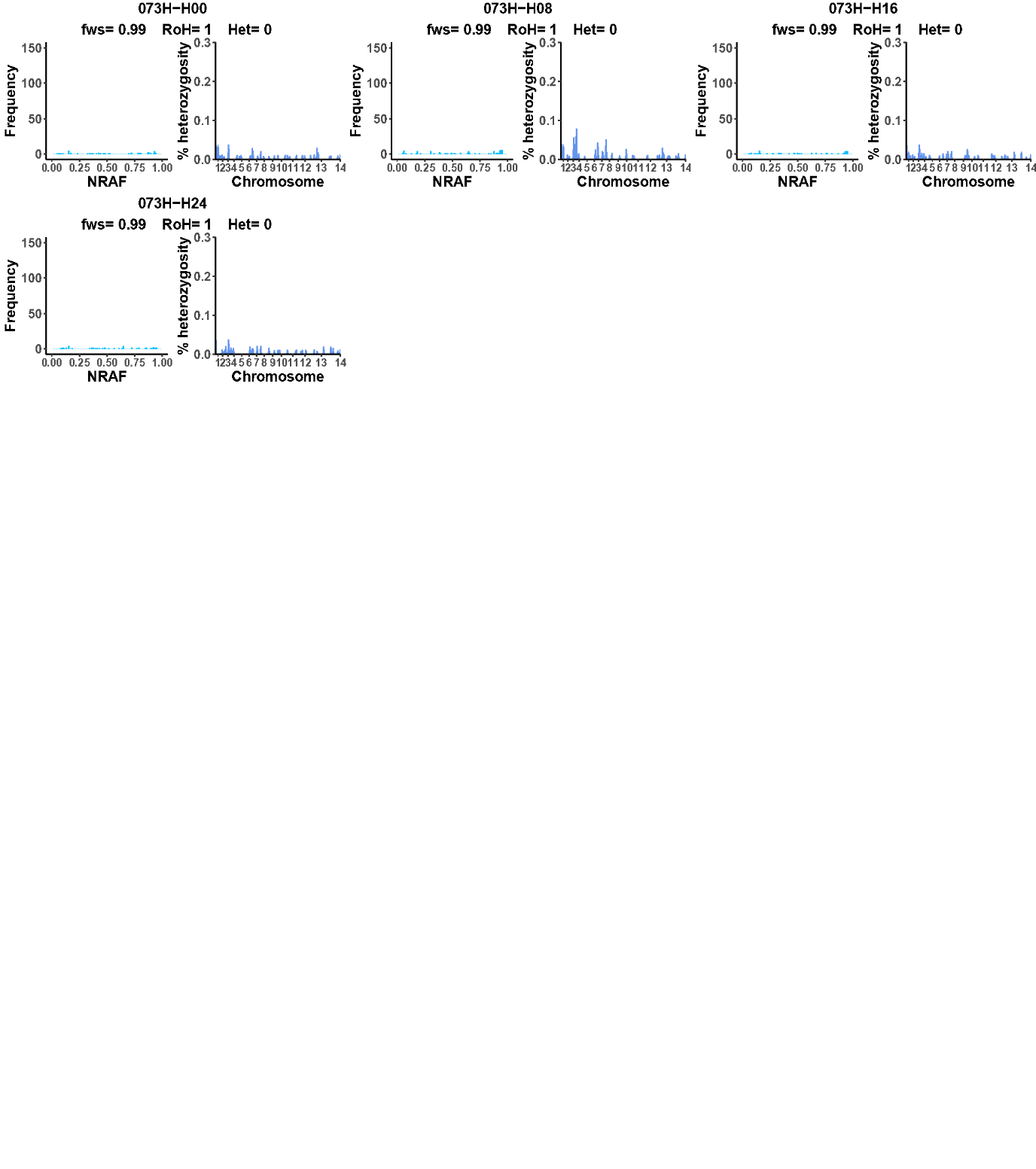

**Supplementary Data 2. Individuals parasite clearance curve by timepoints post-antimalarial treatment. Quinine (A) and artesunate (B) treated groups.**

**A**
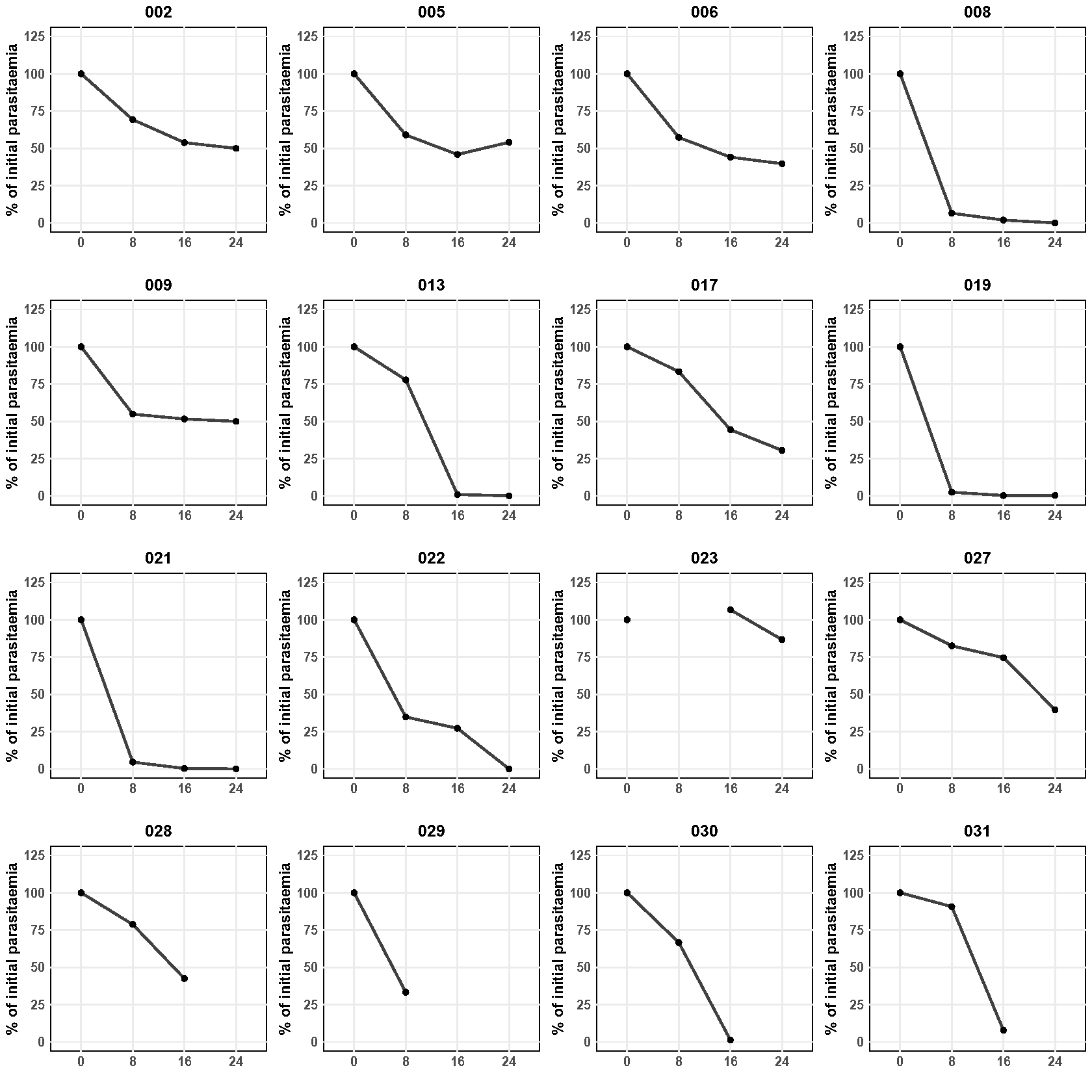

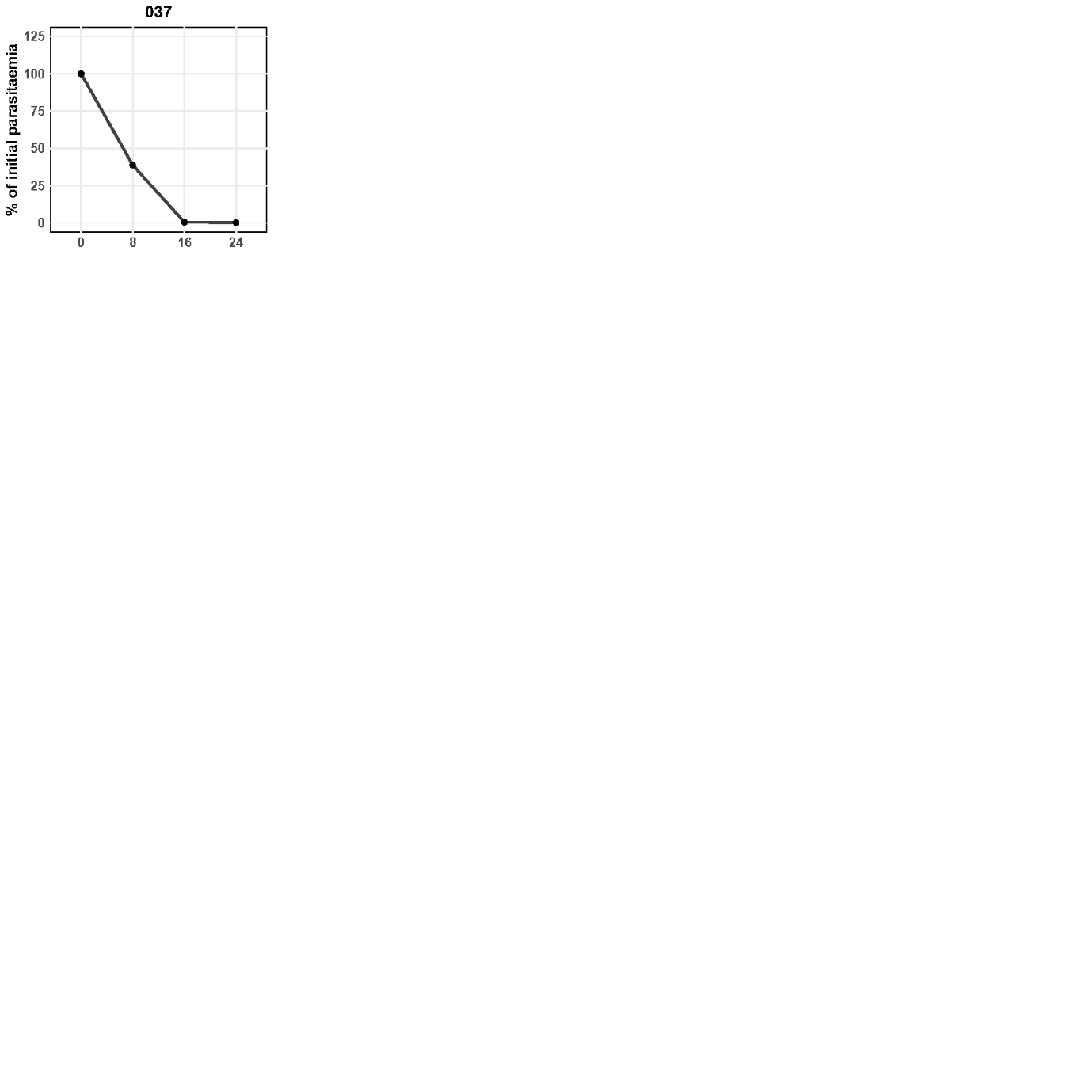

**B**

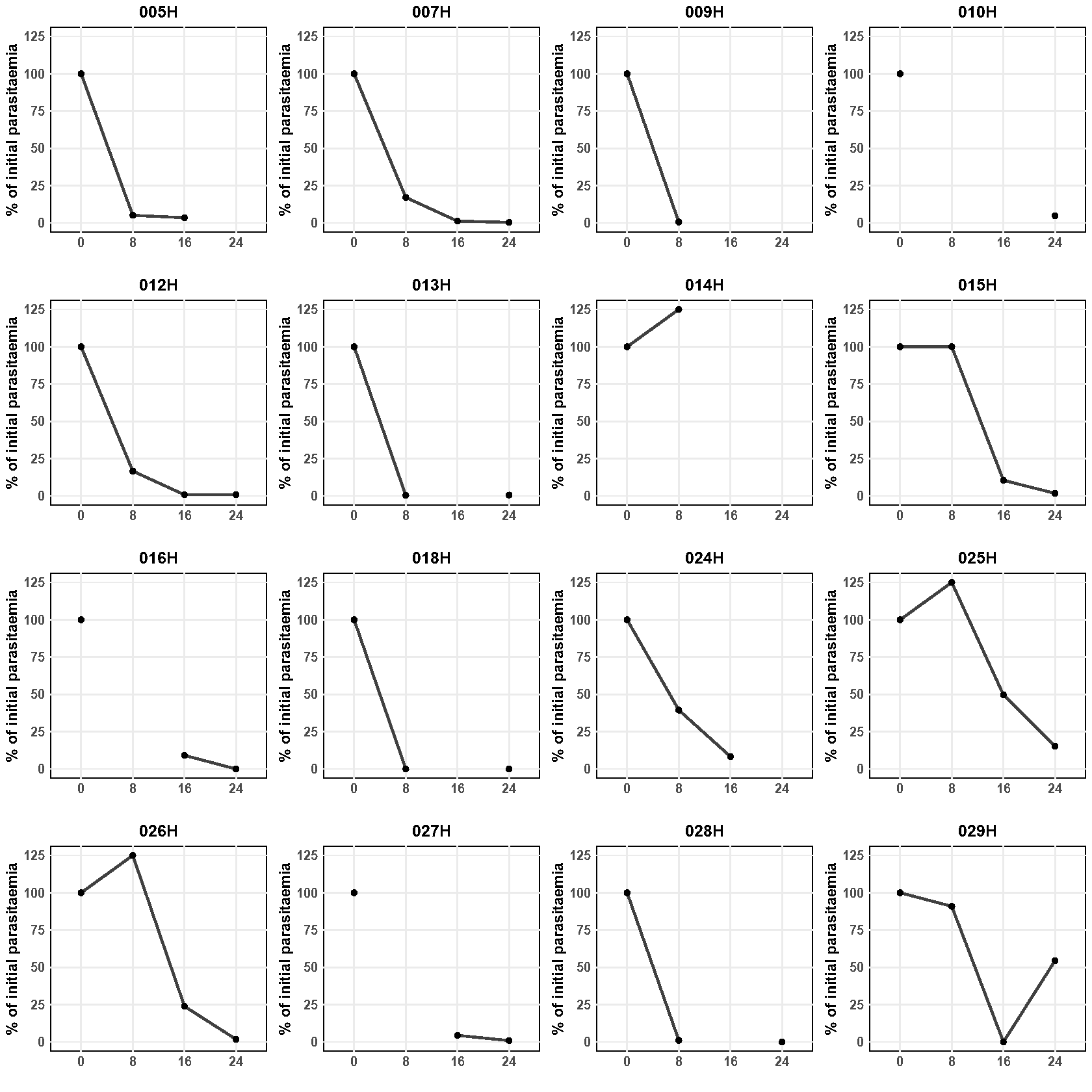

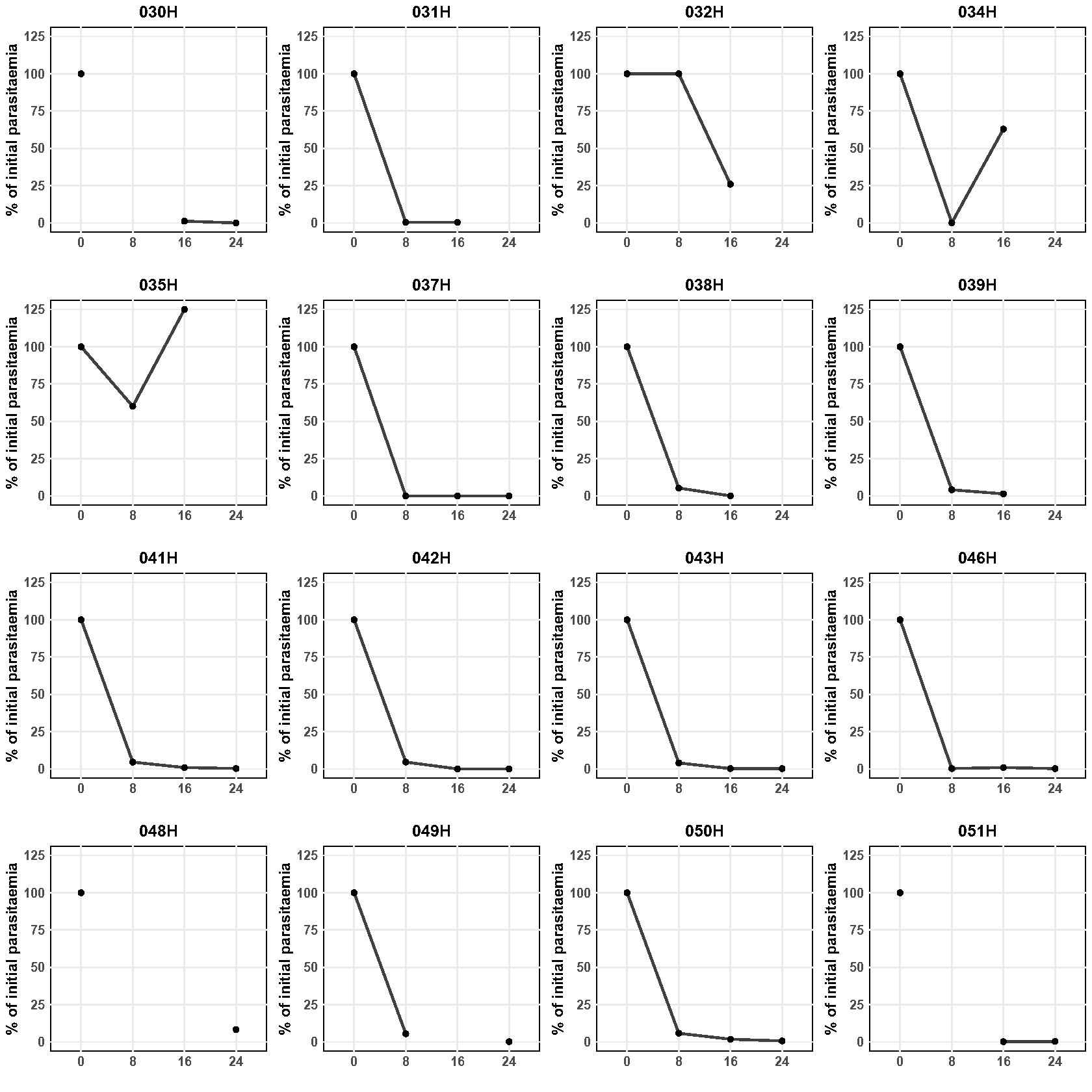

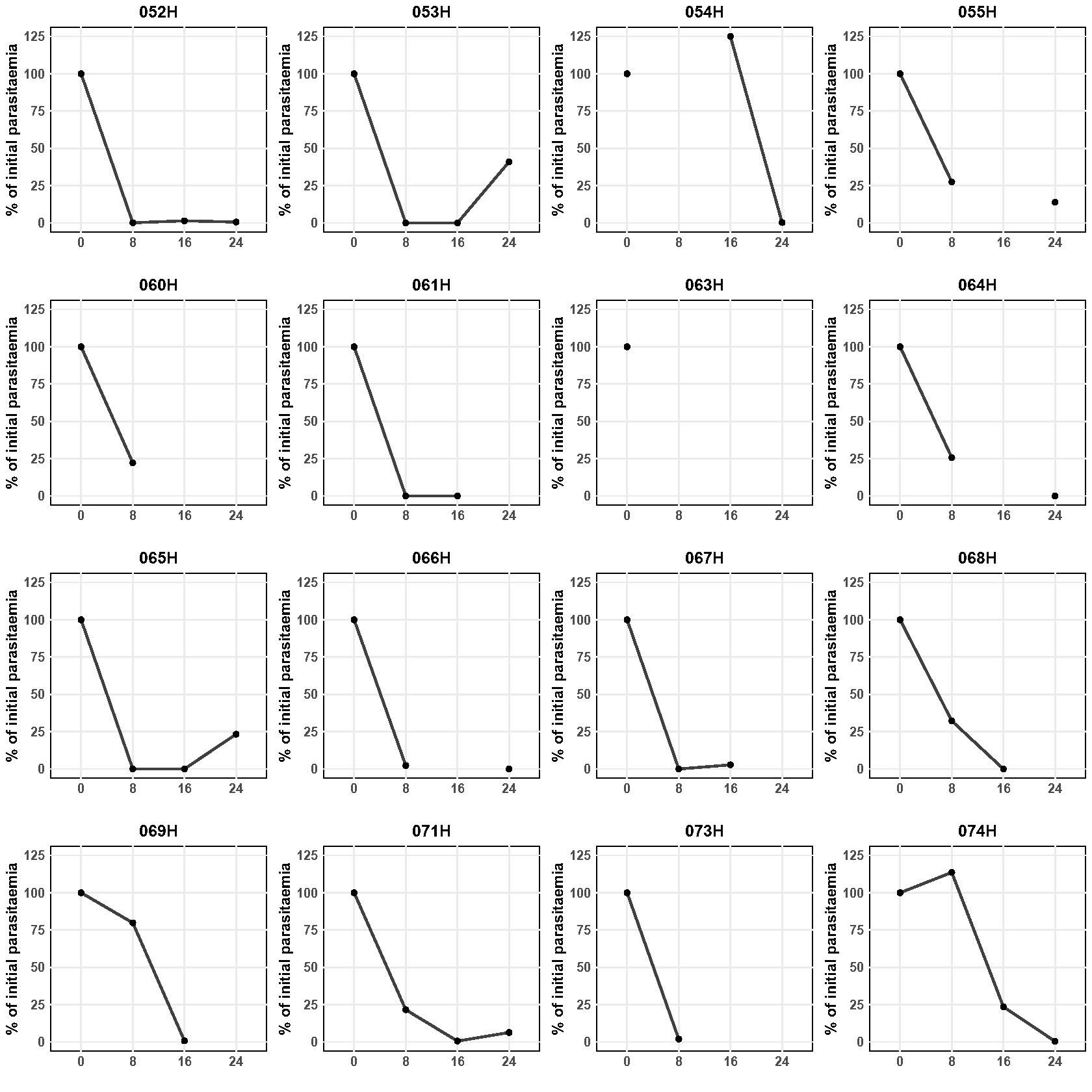
